# SARS-CoV-2 Infections in mRNA Vaccinated Individuals are Biased for Viruses Encoding Spike E484K and Associated with Reduced Infectious Virus Loads that Correlate with Respiratory Antiviral IgG levels

**DOI:** 10.1101/2021.07.05.21259105

**Authors:** Heba H. Mostafa, Chun Huai Luo, C. Paul Morris, Maggie Li, Nicholas J. Swanson, Adannaya Amadi, Nicholas Gallagher, Andrew Pekosz

## Abstract

**Introduction:** COVID-19 large scale immunization in the US has been associated with infrequent breakthrough positive molecular testing. Whether a positive test is associated with a high viral RNA load, specific viral variant, recovery of infectious virus, or symptomatic infection is largely not known.

**Methods:** In this study, we identified 133 SARS-CoV-2 positive patients who had received two doses of either Pfizer-BioNTech (BNT162b2) or Moderna (mRNA-1273) vaccines, the 2nd of which was received between January and April of 2021. The positive samples were collected between January and May of 2021 with a time that extended from 2 to 100 days after the second dose. Samples were sequenced to characterize the whole genome and Spike protein changes and cycle thresholds that reflect viral loads were determined using a single molecular assay. Local SARS-CoV-2 IgG antibodies were examined using ELISA and specimens were grown on cell culture to assess the recovery of infectious virus as compared to a control unvaccinated cohort from a matched time frame.

**Results:** Of 133 specimens, 24 failed sequencing and yielded a negative or very low viral load on the repeat PCR. Of 109 specimens that were used for further genome analysis, 68 (62.4%) were from symptomatic infections, 11 (10.1%) were admitted for COVID-19, and 2 (1.8%) required ICU admission with no associated mortality. The predominant virus variant was the alpha (B.1.1.7), however a significant association between lineage B.1.526 and amino acid change S: E484K with positives after vaccination was noted when genomes were compared to a large control cohort from a matched time frame. A significant reduction of the recovery of infectious virus on cell culture as well as delayed time to the first appearance of cytopathic effect was accompanied by an increase in local IgG levels in respiratory samples of vaccinated individuals but upper respiratory tract IgG levels were not different between symptomatic or asymptomatic infections.

**Conclusions:** Vaccination reduces the recovery of infectious virus in breakthrough infections accompanied by an increase in upper respiratory tract local immune responses.

**Funding:** National Institute of Health (The Johns Hopkins Center of Excellence in Influenza Research and Surveillance, HHSN272201400007C), Johns Hopkins University, Maryland Department of Health, Centers for Disease Control and Prevention.

## Introduction

SARS-CoV-2 has caused a devastating pandemic. Millions of global deaths have been recorded with thousands of new cases diagnosed daily, a trend that significantly changed with the large-scale vaccination in certain countries including the US (https://coronavirus.jhu.edu/map.html). Even though vaccines currently used have high efficacy (1, 2) and undoubtedly, have reduced COVID-19 mortality and severe disease in countries that accelerated mass immunization (3), breakthrough infections have been reported. As of June 1st 2021, the Centers for Disease Control and Prevention (CDC) reported that more than 135 million have been fully vaccinated in the US but so far, the CDC was only notified of 3,016 cases of breakthrough infections that required hospitalization or were associated with mortality (https://www.cdc.gov/vaccines/covid-19/health-departments/breakthrough-cases.html). The frequency of asymptomatic infections though might be underestimated and the relationship between the immune status, viral loads, and recovery of infectious virus from vaccinated positives are largely not known. With the appearance of SARS-CoV-2 variants that are more transmissible or capable of evading vaccine induced immune responses, surveillance has become of utmost importance and genome characterization of positives after vaccination is essential. Currently, data support that vaccines approved for use in the US are effective against most of the currently circulating variants (4)(5). With the general increase in the circulation of variants of concern, it is expected to see a high percentage of breakthrough infections caused by these variants. In our laboratory, as a part of high throughput sequencing for surveillance, positives after full vaccination are genotyped. In addition, viruses are characterized on cell culture to determine the association of virus genotypes and RNA loads with the recovery of infectious virus. In this manuscript, we provide a comprehensive analysis of 133 positives after vaccination diagnosed by Johns Hopkins Clinical Microbiology laboratory. Samples were enrolled in our whole genome sequencing for surveillance pipeline and were retested by the PerkinElmer PCR assay to obtain comparable cycle threshold values (Cts). The recovery of infectious virus from positives after vaccination was determined as well as local SARS-CoV-2 IgG levels in the respiratory samples using ELISA and compared to a control unvaccinated cohort.

## Methods

### Ethical considerations and Data availability

The research Johns Hopkins Medical Institutions Institutional Review Board-X (JHM IRB-X) is constituted to meet the requirements of the Privacy Rule at section 45 CFR 164.512(i)(1)(i)(B) and is authorized and qualified to serve as the Privacy Board for human subjects research applications conducted by Johns Hopkins University faculty members. JHM IRB-3 approved IRB00221396 entitled “Genomic evolution of viral pathogens: impact on clinical severity and molecular diagnosis”. IRB review included the granting of a waiver of consent based on the following criteria: 1) the research involves no more than minimal risk to subjects; 2) the waiver will not adversely affect the rights and welfare of the subjects; 3) the research could not be practicably carried out without the waiver; and 4) the IRB will advise if it is appropriate for participants to be provided with additional pertinent information after participation. This study was also approved for the inclusion of children as ‘research not involving greater than minimal risk’. The permission of parents/guardians is waived. Assent is waived for all children. JHM IRB-X determined that there is no requirement for continuing review or progress report for this application. Remnant nasopharyngeal or lateral mid-turbinate nasal (NMT) clinical swab specimens from patients who tested positive for SARS-CoV-2 after the standard of care testing were used.

### Nucleic acid extraction, PCR, and whole genome sequencing

Automated nucleic acid extraction was performed using the chemagic 360 (PerkinElmer) following the manufacturer’s protocol, with an RNA elution volume of 60µL. Real-time reverse transcriptase PCR (rRT-PCR) was performed using the PerkinElmer SARS-CoV-2 Real-time RT-PCR Assay following the package insert (https://www.fda.gov/media/136410/download). Libraries were prepared in 96 well plates using the ARTIC protocol as previously described (6) with additional clean-up steps using Mag-bind beads prior to combining samples into a single library. Sequencing was performed using the Oxford Nanopore GridION and reads were basecalled with MinKNOW and demultiplexed with guppybarcoder. Reads were size restricted, and alignment and variant calling were performed with the artic-ncov2019 medaka protocol. Clades were determined using Nextclade beta v 0.12.0 (clades.nextstrain.org) (7), and lineages were determined with Pangolin COVID-19 lineage Assigner (COG-UK (cog-uk.io)).

### ELISA

The EUROIMMUN Anti-SARS-CoV-2 ELISA (IgG) was run using undiluted respiratory samples and following the package insert (https://www.fda.gov/media/137609/download). This assay detects antibodies to the S1 domain of the spike protein of SARS-CoV-2. The assay has a cut-off < 0.8 for negative results and ≥ 0.8 to < 1.1 as borderline, which we used as a cut off for nasopharyngeal/ NMT swab specimen types even though these sources were not tested by the manufacturer.

### Cell culture

Vero-TMPRSS2 cells were cultured and infected with aliquots of swab specimens as previously described for VeroE6 cells (8). The presence of SARS-CoV-2 was confirmed by reverse transcriptase PCR (qPCR).

### Statistical analyses

Statistics were performed using GraphPad Prism. For Lineage and Spike analyses Samples with ≥ 90% genome coverage were selected from both vaccinated and control groups (N = 67 for the vaccinated group, and 335 for the control group, Supplementary tables 1 and 3). Controls were selected from samples previously submitted by our group to GISAID, using MatchIt in R (method= ‘optimal’, ratio=5) based on collection date. Control and vaccinated samples were plotted over time to verify a good match. Percentage of samples that matched to lineages were plotted. The full set of mutations present within the spike protein of vaccinated patients were determined, and a heatmap of percentage of samples from vaccinated and control groups was plotted. Chi-square analysis of lineages with at least 5 samples showed a correlation between lineage and vaccine status (*p* = 0.006). Chi-square analysis was performed for lineages P.1, B.1.1.7, B.1.351, B.1.526, and B.1.526.1.

**Table 1.** Clinical and metadata of breakthrough infection cases.

|  | **Positives after full vaccination** | **Positives after full vaccination**<br>**(viral load below the limit of**<br>**detection or false positives)** |
| --- | --- | --- |
| **Total number of patients** | **109** | **24** |
| Median age in years (range) | 51 (23 - >90) | 52 (23 - 79) |
| **No. (%) of** |  |  |
| Male | 40 (36.7) | 6 (25) |
| Female | 69 (63.3) | 18 (75) |
| Symptomatic | 68 (62.4) | 8 (28.6) |
| Asymptomatic | 41 (37.6) | 16 (66.6) |
| **Severity index (%):** |  |  |
| Outpatient | 95 (87.2) | 24 (100) |
| Hospitalized for COVID-19 | 11 (10.1) | 0 |
| ICU admission | 2 (1.8) | 0 |
| **Past Medical History**<br>**Information (%)** |  |  |
| Obesity (BMI > 30) | 18 (16.5) | 3 (12.5) |
| Hypertension | 13 (11.9) | 6 (25) |
| Cardiovascular disease | 13 (11.9) | 1 (4.2) |
| Diabetes | 17 (15.6) | 4 (16.7) |
| Asthma/ chronic lung disease | 16 (14.7) | 5 (20.8) |
| Kidney disease | 11 (10.1) | 2 (8.3) |
| History of cancer/<br>autoimmune disease/ HIV/<br>potentially<br>immunocompromised | 13 (11.9) | 1 (4.2) |
| Median days after COVID-19<br>vaccine (range) | 52 (2 - 99) | 19.5 (3 - 100) |

## Results

### SARS-CoV-2 genomes of positives after full vaccination

As a part of whole genome sequencing of SARS-CoV-2 for surveillance, we identified 133 positives after the completion of either Moderna or Pfizer vaccination regimes that were collected in the time frame of January 2021-May 2021 (Table 1 summarizes the clinical and metadata of this cohort). Of the 133 samples, 24 had failed sequencing with 0% coverage which we believe was related to very low viral loads or false positivity (Supplementary table 1). Twenty-one sequences had coverage less than 50% and average depth of less than 50 and a lineage was not called. Of the genomes that had more than 50% coverage and more than an average depth of 50 (a total of 88), the majority (61%) belonged to the B.1.1.7 (alpha variant) lineage (20I/501Y.V1 clade) followed by the 20C lineages (iota variants) B.1.526 (9%) and B.1.526.1 (4.5%) (Figure 1A and B). A significant correlation between genome coverage and days of sample collection in relation to the second dose of vaccination was noted when all samples with coverage > 1% were included in this analysis (109 total, Supplementary table 1 and Figure 1C, Linear regression, p = 0.049). To correlate genome coverage with the clinical assay’s Ct values, samples were rerun by one diagnostic assay (PerkinElmer) to obtain comparable Ct values. Of the 109 samples with coverage > 1%, 92 specimens had sufficient left-over volumes and were retrieved for rRT-PCR. A significant correlation between Ct values and % coverage was noted with a complete or nearly complete genome coverage associated with Ct values below 25 (Figure 1D, Linear regression, *p* < 0.0001).

**Figure 1.**
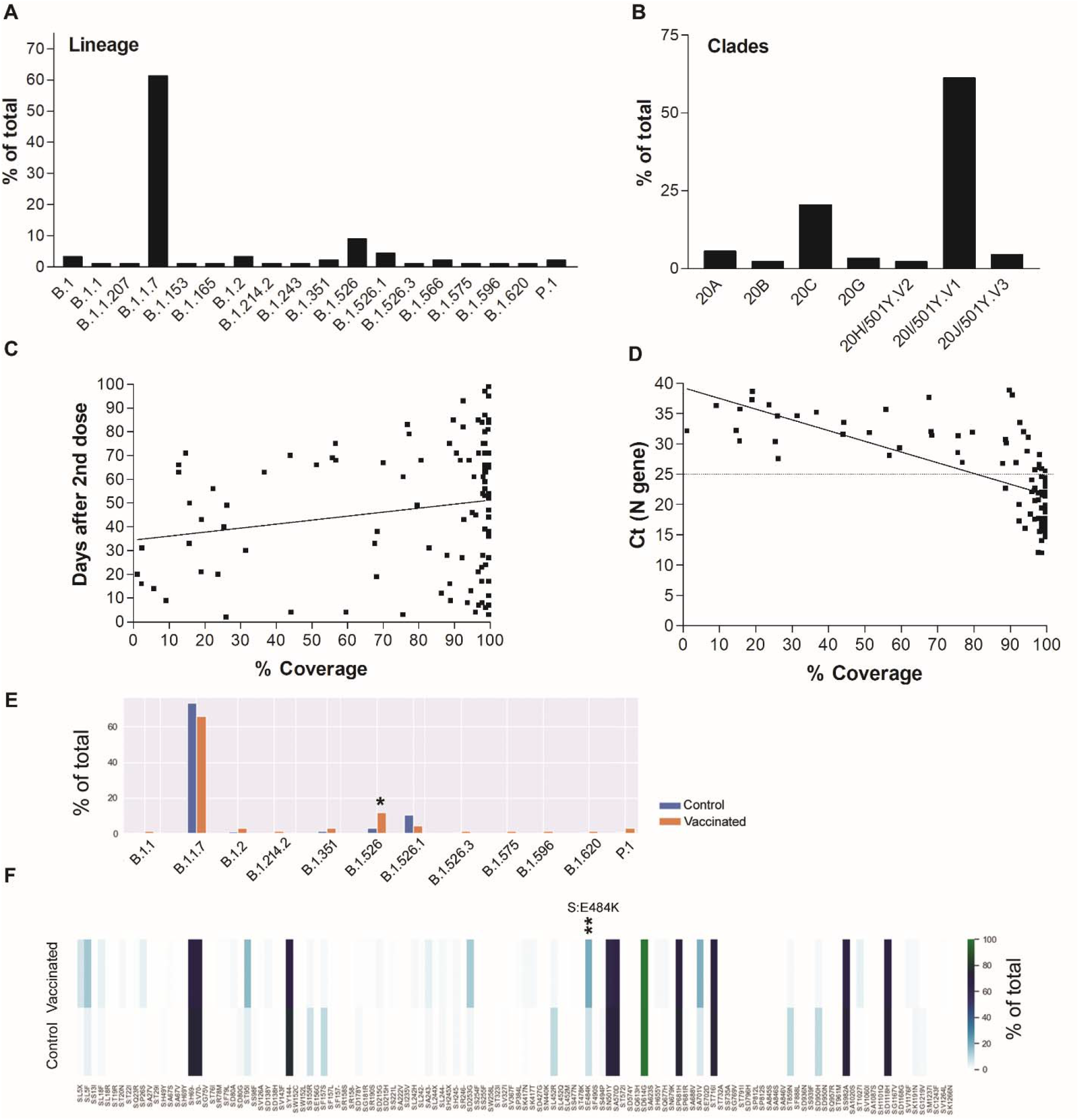
SARS-CoV-2 genomes of positives after full vaccination. A) Lineages and B) Clades of genomes with more than 50% coverage and average depth of 50 (n=88). C) A correlation of SARS-CoV-2 genome coverage with the days of sample collection after the second dose of vaccination. D) A correlation between SARS-CoV-2 genome coverage and cycle thresholds of the N gene using the PerkinElmer SARS-CoV-2 assay. E) A comparison between lineages from fully vaccinated (n = 67) and control (n = 335) genomes with coverage ≥ 90%. F) Spike amino acid changes in vaccinated and control groups. * *p* < 0.05, *** *p* < 0.005

When a large cohort of our characterized genomes were randomly selected as a control based on the sample collection dates and only genomes with ≥ 90% coverage were used for the analysis (vaccinated: N = 67, and control: N = 335), an association was seen between B.1.526 and vaccination status (Chi-square with Bonferroni correction *p* = 0.022, Figure 1E). Spike substitution analysis showed that the S: E484K is associated with genomes of the vaccinated group (*p* = 0.0032, Figure 1F and Supplementary tables 3, 4, and 5).

### Recovery of infectious virus from fully vaccinated individuals and the correlation with local SARS-CoV-2 antibodies and viral loads

To assess the recovery of infectious virus from positive samples of fully vaccinated individuals, first, a cohort of control samples was selected. A total of 124 positive samples from unvaccinated individuals collected in the time frame between the end of December 2020 to the first week of March 2021 were used for comparison (Supplementary table 2). The cohort was selected to include the alpha variant lineage as well as other predominant lineages before the month of March. To compare the recovery of infectious virus between positives from fully vaccinated (N = 114 with sufficient volume) and control (N = 124) groups, samples were cultured on Vero-TMPRSS2 cells and the time to cytopathic effect (CPE) was compared between the two groups. As expected, samples that had failed sequencing and were thought of as very low viral load or false positives did not yield infectious virus and were excluded from further analysis (Supplementary table 1). Of the fully vaccinated group, 17 of 92 samples (18.5%) showed CPE on cell culture compared to 80 out of 124 (64.5%) of the control group (Fisher Exact test, *p* < 0.00001). Notably, the control group recovery on cell culture was faster than the vaccinated group with 44 out of 80 samples (55%) positive 2 days after culture compared to no samples showing CPE for the vaccinated group (Figure 2 A and B).

**Figure 2.**
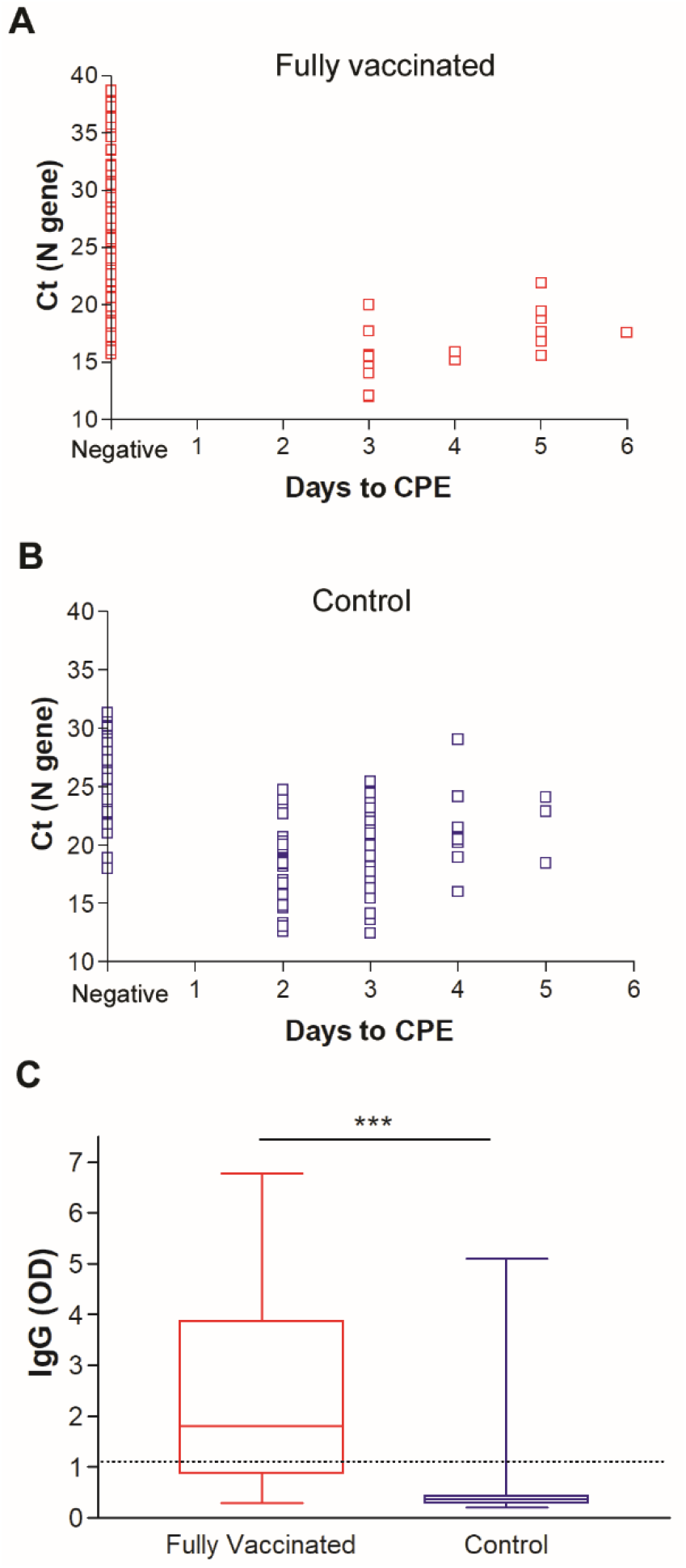
Recovery of infectious SARS-CoV-2 on Vero-TMPRSS2 cells for A) fully vaccinated and B) control groups. C) SARS-CoV-2 IgG in upper respiratory swab samples from fully vaccinated and control groups. Dashed line demarcates the limit of borderline and negative ELISA results as specified per assay’s package insert (1.1). * *p* < 0.05, *** *p* < 0.0001

To study the local SARS-CoV-2 antibodies in the respiratory samples and their correlation to the observed CPE phenotype, respiratory samples grown on cell culture were also tested by ELISA for SARS-CoV-2 IgG. A significant increase in SARS-CoV-2 nasal and nasopharyngeal IgG levels were noted in the respiratory samples from vaccinated individuals when compared to the control group (Figure 2C, *t* test, *P* < 0.0001).

To better understand the contribution of Ct values and local antibodies on virus recovery on cell culture, we focused our analysis on samples with Ct values below 25 which constituted 49 samples in the vaccinated group versus 96 samples in the control group (Supplementary tables 1 and 2). Notably the distribution of the Ct values between the two groups for samples with Ct values lower than 25 was not different (Figure 3A). The majority of the control group samples were positive on cell culture (77, 80.2%), in contrast to 17 (34.7%) of the vaccinated group (Figure 3B). Consistent with data from the whole cohort, higher nasal/ nasopharyngeal IgG levels (Figure 3C, *P* < 0.0001) was noted for the fully vaccinated group.

**Figure 3.**
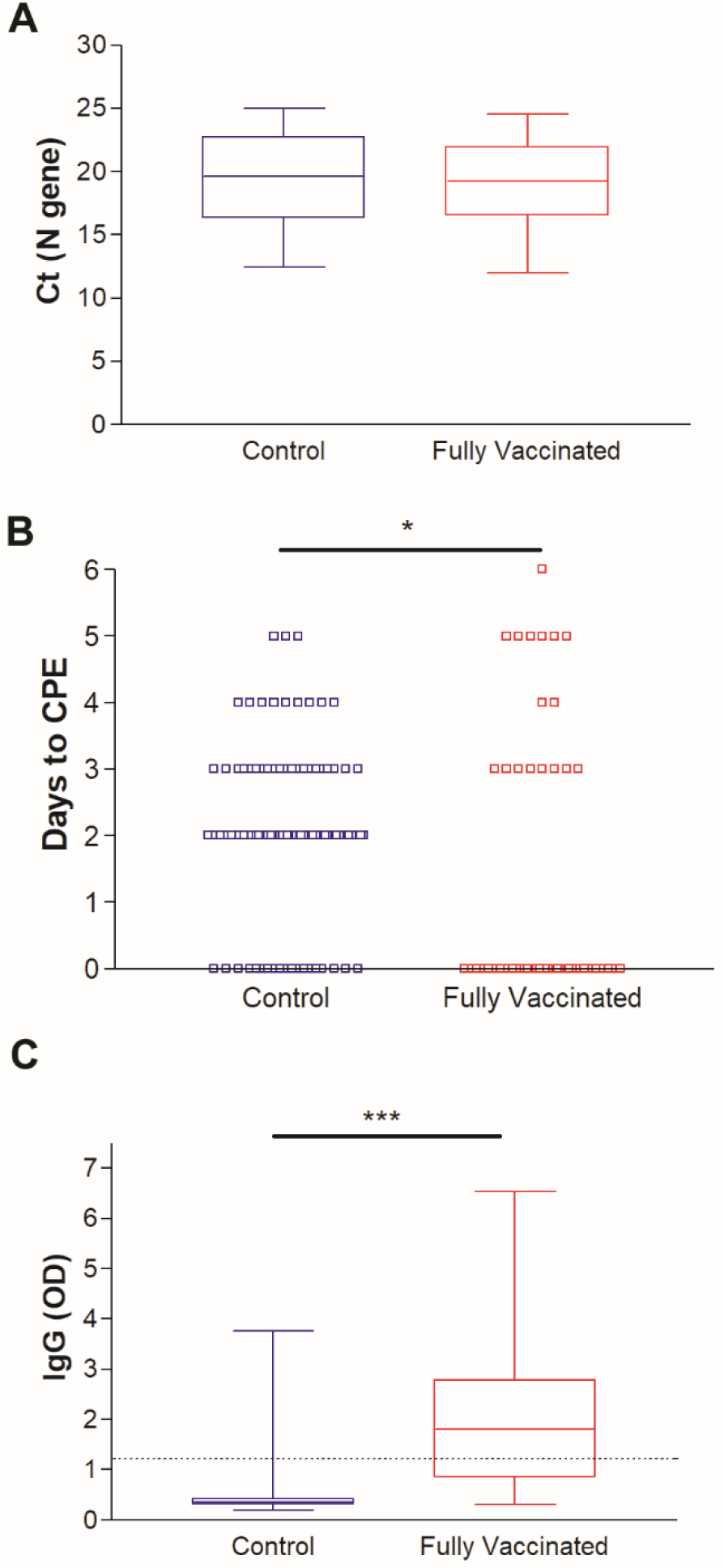
Recovery of infectious SARS-CoV-2 on Vero-TMPRSS2 cells for samples with Ct values less than 25 (N gene). A) comparison of Ct values distribution between control and fully vaccinated groups. B) Days to CPE in control and fully vaccinated groups. C) IgG in vaccinated and control groups respiratory samples. Dashed line demarcates the limit of borderline and negative ELISA results as specified per assay’s package insert (1.1). * *p* < 0.05, *** *p* < 0.0001

### Characterization of local SARS-CoV-2 antibody responses

A correlation of SARS-CoV-2 local IgG with the days of sample collection after receiving the second dose of the COVID-19 vaccine showed a trend of reduction with the progress of time (Figure 4A, linear regression, *p* < 0.0001). No significant correlations between the IgG levels and Ct values were noticed (Figure 4B). Negative recovery of infectious virus on cell culture correlated with high levels of IgG in respiratory specimens, with no infectious virus isolated from samples with an IgG OD reading of >3.0 (Figure 4C, *t* test, *P* = 0.004). The presence of symptoms did not correlate with higher IgG levels when we compared samples from symptomatic vaccinated to asymptomatic vaccinated individuals (Figure 5A). Notably, a significant increase in the mean Ct value for the asymptomatic group was noted (27.6 versus 23.2, *t* test, *P* = 0.0048, Figure 5B) as well as a reduction in the mean genome coverage (70.8% versus 84.9%, *t* test, *P* = 0.0284, Figure 5C). Notably, infectious virus was recovered from only 2 samples from asymptomatic patients (6.5%) in contrast to 15 from symptomatic patients (24.6%) (Supplementary table 1).

**Figure 4.**
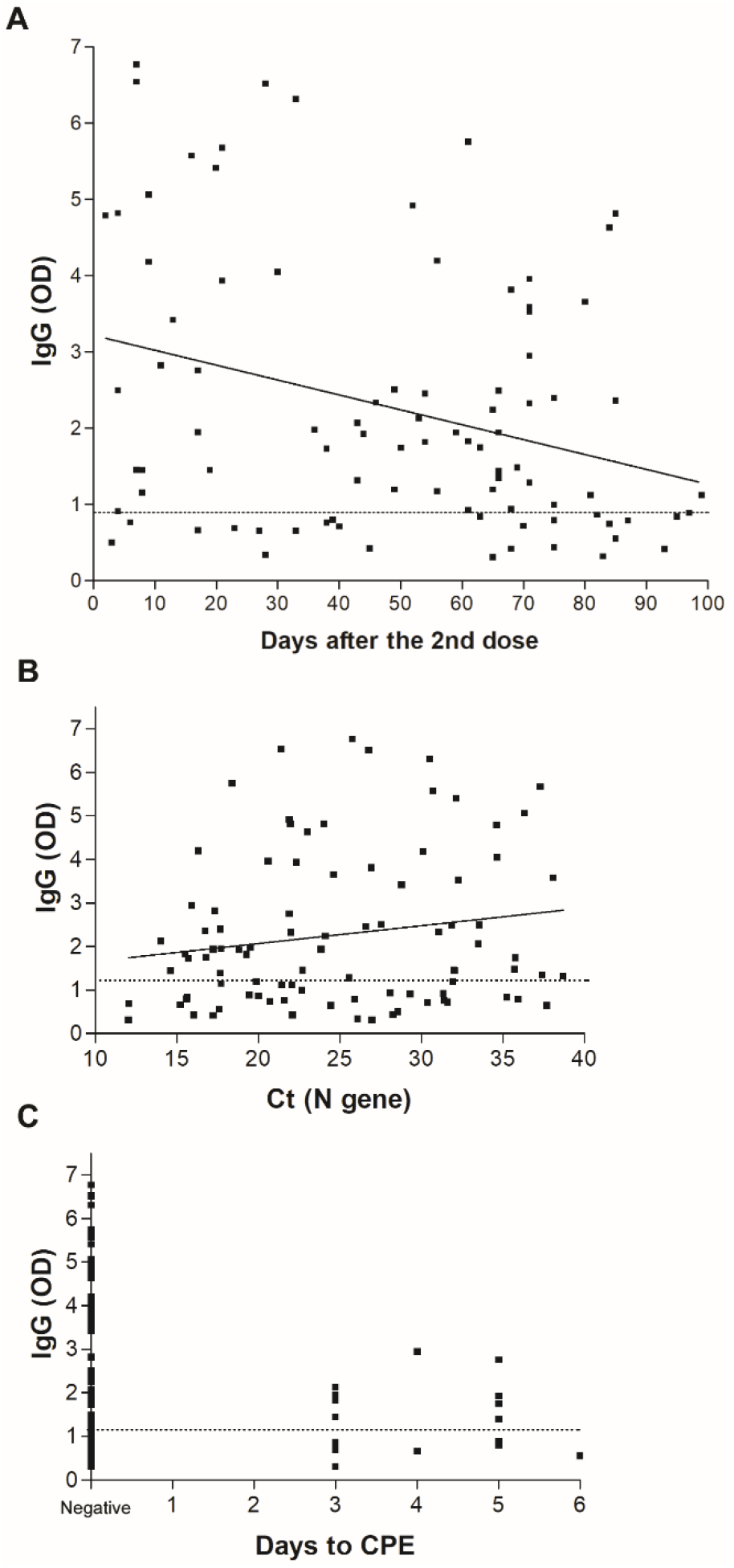
Local SARS-CoV-2 antibodies in upper respiratory samples of vaccinated individuals. A) IgG levels by ELISA in the upper respiratory samples collected from patients positive after full vaccination (N=114) and association with the days after receiving the second dose of the COVID-19 vaccine. B) SARS-CoV-2 IgG correlation to cycle thresholds of the N gene using the PerkinElmer SARS-CoV-2 assay. C) SARS-CoV-2 IgG correlation to days to the first appearance of cytopathic effect (CPE) on Vero-TMPRSS2 cells. Dashed line demarcates the limit of borderline and negative ELISA results as specified per assay’s package insert.

**Figure 5.**
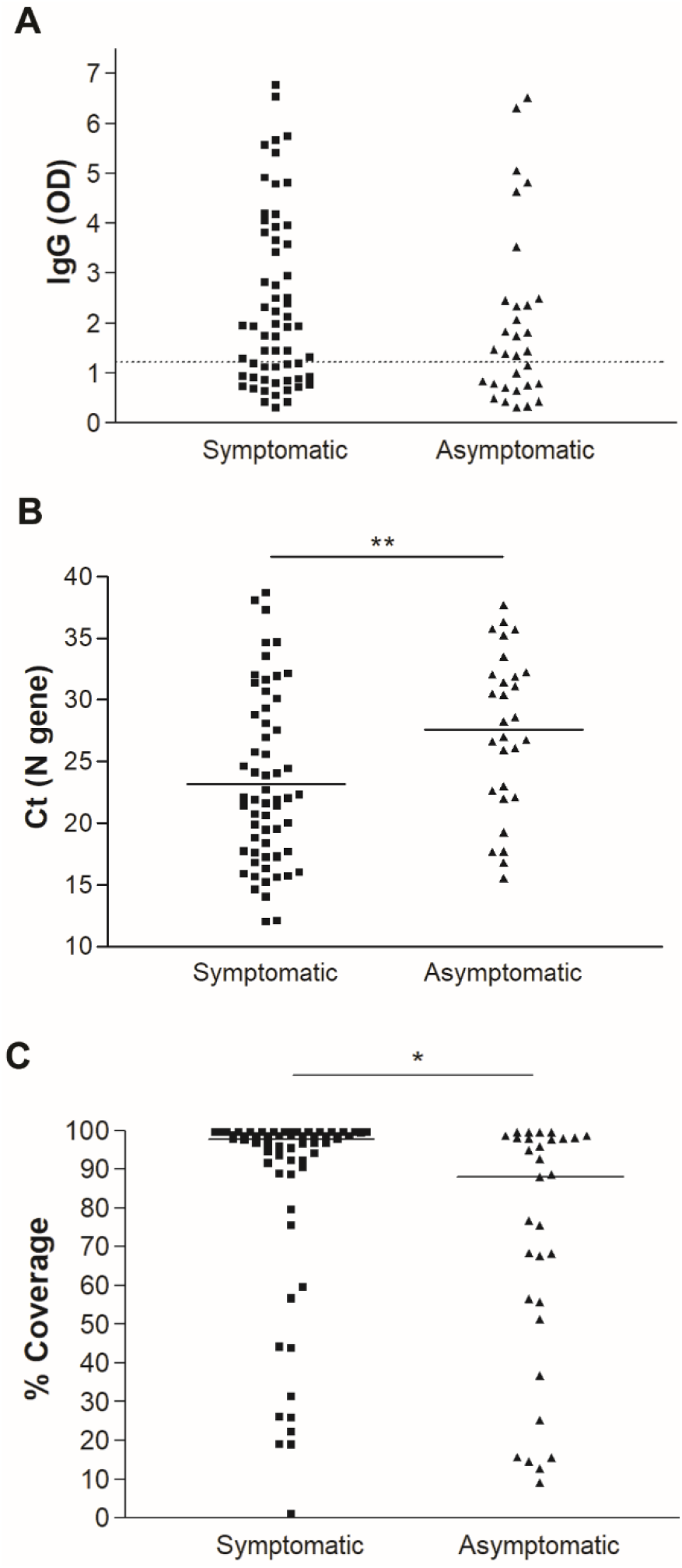
Local antibodies, viral loads, and recovery of whole genomes in symptomatic versus asymptomatic vaccinated individuals. A) IgG levels by ELISA in the upper respiratory samples. B) Comparison of Ct values distribution between symptomatic and asymptomatic groups. C) % genome coverage in symptomatic and asymptomatic groups. Dashed line demarcates the limit of borderline and negative ELISA results as specified per assay’s package insert. * *p* < 0.05, ** *p* < 0.005

## Discussion

In this study, we provide a comprehensive analysis on a cohort of 133 SARS-CoV-2 positive specimens collected after the completion of COVID-19 vaccination. This cohort was enrolled in our whole genome sequencing for SARS-CoV-2 surveillance. Genomic analysis revealed that only 67 had genomic coverage of ≥ 90% and when compared to a random cohort of our characterized genomes from a matched time frame, there was a statistically significant increased representation of lineage B.1.526 as well as the Spike amino acid change S:E484K. Strikingly, when samples with Ct values less than 25 were compared to a control cohort of similarly distributed Ct values, the recovery of infectious virus from cases post-vaccination was significantly impaired, evident as both a delay in the first appearance of cytopathic effect as well as a significant decrease in the total number of positive samples on cell culture. This data indicates that infection in vaccinated individuals results in reduced infectious virus load compared to unvaccinated individuals and that the Ct value from infected, vaccinated individuals is associated with lower infectious virus loads compared to unvaccinated individuals. This may further reduce the likelihood that infected, vaccinated individuals can transmit SARS-CoV-2 to others. Interestingly, the lower infectious virus load in vaccinated individuals was associated with an increase in local SARS-CoV-2 specific IgG levels. The reduction in infectious virus in samples with Ct values of less than 25 could be explained if the nasal SARS-CoV-2 IgGs are neutralizing. The recovery of infectious virus was higher in symptomatic when compared to asymptomatic vaccinated individuals, but local SARS-CoV-2 IgG levels were comparable between symptomatic and asymptomatic cases, indicating that local antiviral IgG levels do not drive higher asymptomatic infection rates. Recent data from the CDC showed that the widely used mRNA vaccines in the US reduce the infection risk by 91% and data from different groups confirm that breakthrough infections after full vaccination are scarce (9-11). Data also show that vaccines reduce symptomatic and asymptomatic infections (12-14). Our data is consistent with published data, and even though the total number of infections after vaccination is not known, the occurrence of positives after full vaccination in our surveillance cohort has been infrequent. Our data show that the 18% of 133 positive individuals who tested positives after the second dose were likely false positives or had very low viral loads that rendered the repeat rRT-PCR runs and sequencing negatives. Our data also show that 37.6 % of these breakthrough infections were asymptomatic and in symptomatic patients, the majority were mild cases that did not require hospitalization.

The correlation between positive PCR results and the recovery of infectious virus after COVID-19 vaccination was not previously reported, however in vivo animal studies showed that vaccines reduce viral replication in the respiratory tracts of animals (15-17). Vaccines were also shown to reduce SARS-CoV-2 transmission and a recent report showed a reduction in viral RNA loads after vaccination (18). Our data show that, despite a similar distribution of Ct values in vaccinated and control unvaccinated individuals, the recovery of infectious virus after vaccination is significantly attenuated. The recovery of infectious virus though could still infrequently happen and hence might be associated with reduced transmission. Additional studies are required to determine the kinetics of infectious virus shedding in vaccinated individuals.

The emergence of SARS-CoV-2 variants of concern and interest were associated with changes in the spike protein within regions that could affect the receptor binding domain or impact the neutralization of the virus by natural or vaccine induced immune responses (19-22). Those variants were associated with an increase in transmissibility and in particular the S: E484K substitution was associated with a compromise in the neutralization by monoclonal antibodies rendering this change “of therapeutic concern”. The S: E484K independently emerged in multiple lineages in distant geographical locations including the B.1.351 and the P.1 and those lineages showed some reduction in neutralization by sera collected from immunized individuals as well as decreased susceptibility to certain therapeutic monoclonal antibodies. Additionally, the B.1.351 and P.1 were associated with reductions in the vaccine efficacy data in locations of its predominance (23, 24). The S: E484K is also present in some strains of lineage B.1.526, a lineage which was significantly associated with positives after vaccination in our cohort, even though in a previous study, it was not reported to associate with positives after vaccination (25). Our study shows that the S: E484K is significantly associated with breakthrough cases after vaccination in a well-controlled analysis that used a large cohort of controls from a matched time frame of sample collection. This data highlights the significance of continuous genomic surveillance coupled with metadata and laboratory data collections for providing significant information with the potential to impact decisions related to booster doses or modifying vaccine strains.

In conclusion, this is the first study that combines genomic analysis, cell culture, and mucosal serology to correlate reduced recovery of infectious virus from positives after vaccination with increased local IgG levels. This study is also the first to show the significant association of S: E484K with positives after full vaccination using a well-controlled analysis and a relatively large sample size.

## Data Availability

All data are available within the manuscript and in the supplemental tables.

## Data sharing

Whole genome data were made available publicly and raw genomic data requests could be directed to HHM.

## Acknowledgement

This study was only possible with the unique efforts of the Johns Hopkins Clinical Microbiology Laboratory faculty and staff. We also thank the Johns Hopkins Immunology Laboratory and Kim Jeemin for technical assistance. HHM is supported by the HIV Prevention Trials Network (HPTN) sponsored by the National Institute of Allergy and Infectious Diseases (NIAID), National Institute on Drug Abuse, National Institute of Mental Health, and Office of AIDS Research, of the NIH, DHHS (UM1 AI068613), the NIH RADx-Tech program (3U54HL143541-02S2), National Institute of Health RADx-UP initiative (Grant R01 DA045556-04S1), National Institute of Allergy and Infectious Diseases (Johns Hopkins Center of Excellence in Influenza Research and Surveillance HHSN272201400007C), the U. S. Centers for Disease Control (75D30121C11061), Johns Hopkins University President’s Fund Research Response, the Johns Hopkins Department of Pathology, and the Maryland Department of Health. This research was supported in part by the intramural research program of the National Institutes of Health. The views expressed in this manuscript are those of the authors and do not necessarily represent the views of the National Institute of Biomedical Imaging and Bioengineering; the National Heart, Lung, and Blood Institute; the National Institutes of Health, the U.S. Centers for Disease Control or the U.S. Department of Health and Human Services.

