## supplemental tables for "SARS-CoV-2 Infections in mRNA Vaccinated Individuals are Biased for Viruses Encoding Spike E484K and Associated with Reduced Infectious Virus Loads that Correlate with Respiratory Antiviral IgG levels"

Suppl. Table 1

| HPID | Clade | Lineage | % coverage | Depth | IgG (OD) | E ORF1ab ( | PE N Ct | ter 2nd Vacci | Symptoms | Days to CPE |
|---|---|---|---|---|---|---|---|---|---|---|
| HP04818 | 20I/501Y.V1 | B.1.1.7 | 99.60 | 426.21 | 2.36 | 22.88 | 16.75 | 85 | no | negative |
| HP05815 | 20I/501Y.V1 | B.1.1.7 | 99.60 | 428.57 | 1.12 | 22.62 | 22.08 | 99 | yes | negative |
| HP05816 | 20C | B.1.526 | 99.60 | 429.57 | 6.54 | 21.25 | 21.41 | 7 | yes | negative |
| HP03729 | 20I/501Y.V1 | B.1.1.7 | 99.60 | 445.19 | 1.73 | 16.74 | 15.73 | 38 | yes | negative |
| HP03775 | 20I/501Y.V1 | B.1.1.7 | 99.60 | 449.04 | 1.92 | 18.74 | 18.82 | 44 | yes | 5 |
| HP03786 | 20I/501Y.V1 | B.1.1.7 | 99.60 | 392.08 | 1.82 | 19.12 | 19.27 | 54 | no | negative |
| HP04129 | 20I/501Y.V1 | B.1.1.7 | 99.60 | 443.65 | 2.24 | 24.22 | 24.12 | 65 | yes | negative |
| HP04153 | 20I/501Y.V1 | B.1.1.7 | 99.60 | 447.43 | 1.98 | 19.82 | 19.51 | 36 | yes | negative |
| HP04182 | 20I/501Y.V1 | B.1.1.7 | 99.60 | 438.17 | 0.65 | 24.47 | 24.44 | 27 | yes | negative |
| HP04113 | 20C | B.1.526 | 99.60 | 446.86 | 1.20 | 20.02 | 19.87 | 65 | yes | negative |
| HP04414 | 20I/501Y.V1 | B.1.1.7 | 99.60 | 427.64 | 1.44 | 15.97 | 14.60 | 66 | yes | 3 |
| HP04499 | 20I/501Y.V1 | B.1.1.7 | 99.60 | 415.81 | 4.63 | 23.18 | 23.00 | 84 | no | negative |
| HP04923 | 20G | B.1.2 | 99.60 | 436.58 | Not done | Not done | Not done | 47 | yes | negative |
| HP04930 | 20I/501Y.V1 | B.1.1.7 | 99.60 | 441.61 | Not done | Not done | Not done | 7 | yes | negative |
| HP04931 | 20I/501Y.V1 | B.1.1.7 | 99.60 | 436.61 | Not done | Not done | Not done | 3 | yes | negative |
| HP04957 | 20I/501Y.V1 | B.1.1.7 | 99.60 | 397.38 | 1.94 | 25.25 | 23.86 | 66 | yes | negative |
| HP05000 | 20C | B.1.526 | 99.60 | 431.83 | 1.95 | 17.42 | 17.70 | 17 | yes | 3 |
| HP05016 | 20I/501Y.V1 | B.1.1.7 | 99.60 | 366.65 | 1.28 | 27.58 | 25.56 | 71 | yes | negative |
| HP05024 | 20I/501Y.V1 | B.1.1.7 | 99.60 | 418.68 | 4.92 | 22.73 | 21.89 | 52 | yes | negative |
| HP05279 | 20I/501Y.V1 | B.1.1.7 | 99.60 | 433.09 | 2.82 | 18.42 | 17.34 | 11 | Yes | negative |
| HP05604 | 20C | B.1.526 | 99.60 | 442.55 | 0.66 | 15.20 | 15.21 | 17 | Yes | 4 |
| HP04481 | 20J/501Y.V3 | P.1 | 99.59 | 437.41 | 0.84 | 16.59 | 15.66 | 95 | yes | 3 |
| HP05031 | 20I/501Y.V1 | B.1.1.7 | 99.59 | 437.20 | 4.81 | 22.41 | 21.98 | 85 | No | negative |
| HP03722 | 20C | B.1.526 | 99.59 | 440.68 | 2.95 | 18.37 | 15.92 | 71 | yes | 4 |
| HP04993 | 20A | B.1.214.2 | 99.49 | 426.97 | 1.12 | 21.90 | 21.44 | 81 | yes | negative |
| HP05512 | 20I/501Y.V1 | B.1.1.7 | 98.70 | 265.03 | 0.76 | 21.79 | 21.60 | 6 | Yes | negative |
| HP05616 | 20I/501Y.V1 | B.1.1.7 | 98.70 | 405.92 | Not done | Not done | Not done | 24 | No | negative |
| HP05025 | 20A | B.1.620 | 98.68 | 429.94 | Not done | Not done | Not done | 63 | yes | negative |
| HP03453 | 20I/501Y.V1 | B.1.1.7 | 98.60 | 419.70 | 4.20 | 18.21 | 16.32 | 56 | yes | negative |
| HP04204 | 20I/501Y.V1 | B.1.1.7 | 98.60 | 407.75 | 3.66 | 25.15 | 24.60 | 80 | yes | negative |
| HP04320 | 20I/501Y.V1 | B.1.1.7 | 98.60 | 392.79 | 1.74 | 16.80 | 16.81 | 63 | yes | 5 |
| HP04461 | 20I/501Y.V1 | B.1.1.7 | 98.60 | 418.46 | 1.38 | 17.66 | 17.66 | 66 | no | 5 |

| | | | | | | | | | | |
|---|---|---|---|---|---|---|---|---|---|---|
| HP04464 | 20I/501Y.V1 | B.1.1.7 | 98.60 | 381.40 | 0.79 | 25.68 | 25.91 | 87 | no | negative |
| HP05130 | 20I/501Y.V1 | B.1.1.7 | 98.60 | 420.76 | 1.94 | 17.99 | 17.22 | 59 | Yes | negative |
| HP05159 | 20I/501Y.V1 | B.1.1.7 | 98.60 | 411.48 | 0.80 | 16.17 | 15.61 | 39 | Yes | 5 |
| HP05181 | 20I/501Y.V1 | B.1.1.7 | 98.60 | 410.03 | 0.89 | 20.11 | 19.45 | 97 | Yes | 5 |
| HP05595 | 20I/501Y.V1 | B.1.1.7 | 98.60 | 288.57 | 2.39 | 18.12 | 17.68 | 75 | Yes | negative |
| HP05581 | 20C | B.1.526 | 98.60 | 288.49 | 0.31 | 11.85 | 12.02 | 65 | Yes | 3 |
| HP04826 | 20I/501Y.V1 | B.1.1.7 | 98.60 | 405.38 | 2.32 | 22.03 | 22.00 | 71 | yes | negative |
| HP05129 | 20C | B.1.526 | 98.60 | 404.86 | 2.13 | 14.86 | 14.04 | 53 | Yes | 3 |
| HP05253 | 20C | B.1.575 | 98.05 | 338.42 | 0.43 | 27.08 | 28.25 | 75 | No | negative |
| HP04656 | 20I/501Y.V1 | B.1.1.7 | 98.03 | 350.64 | 1.83 | 16.04 | 15.52 | 61 | no | 3 |
| HP04632 | 20I/501Y.V1 | B.1.1.7 | 97.99 | 323.89 | 2.45 | 27.04 | 26.60 | 54 | no | negative |
| HP05509 | 20H/501Y.V2 | B.1.351 | 97.97 | 287.24 | 0.34 | 24.75 | 26.09 | 28 | No | negative |
| HP05160 | 20I/501Y.V1 | B.1.1.7 | 97.77 | 419.46 | 1.15 | 17.65 | 17.71 | 8 | No | negative |
| HP05201 | 20I/501Y.V1 | B.1.1.7 | 97.75 | 379.48 | 0.74 | 21.75 | 20.73 | 84 | Yes | negative |
| HP04665 | 20C | B.1.526.1 | 97.70 | 429.27 | 0.69 | 12.25 | 12.08 | 23 | yes | 3 |
| HP04198 | 20G | B.1.596 | 97.65 | 397.35 | 2.76 | 23.15 | 21.90 | 17 | yes | 5 |
| HP04430 | 20I/501Y.V1 | B.1.1.7 | 96.78 | 368.56 | 3.96 | 20.93 | 20.61 | 71 | yes | negative |
| HP05168 | 20I/501Y.V1 | B.1.1.7 | 96.76 | 350.76 | 1.45 | 22.54 | 22.71 | 7 | Yes | negative |
| HP05171 | 20I/501Y.V1 | B.1.1.7 | 96.76 | 377.93 | 6.77 | 25.97 | 25.75 | 7 | Yes | negative |
| HP05145 | 20J/501Y.V3 | P.1 | 96.71 | 414.12 | 0.55 | 18.57 | 17.60 | 85 | Yes | 6 |
| HP03053 | 20I/501Y.V1 | B.1.1.7 | 96.62 | 273.11 | 3.93 | 22.45 | 22.32 | 21 | yes | negative |
| HP05105 | 20I/501Y.V1 | B.1.1.7 | 95.98 | 330.81 | 0.42 | 22.18 | 22.09 | 45 | No | negative |
| HP05084 | 20C | B.1.526.1 | 95.88 | 346.84 | 4.82 | 24.13 | 24.04 | 4 | Yes | negative |
| HP05277 | 20C | B.1.526 | 95.49 | 179.36 | 5.75 | 18.11 | 18.40 | 61 | Yes | negative |
| HP04074 | 20I/501Y.V1 | B.1.1.7 | 94.98 | 218.59 | 2.34 | 30.85 | 31.08 | 46 | no | negative |
| HP05805 | 20I/501Y.V1 | B.1.1.7 | 94.64 | 297.75 | 3.42 | 30.86 | 28.80 | 13 | yes | negative |
| HP05624 | 20C | B.1.2 | 94.13 | 96.92 | 0.42 | 16.51 | 16.05 | 68 | Yes | negative |
| HP02169 | 20B | B.1.1 | 93.61 | 154.00 | 1.45 | 31.21 | 32.00 | 8 | yes | negative |
| HP03374 | 20C | B.1.526.1 | 92.62 | 229.55 | 2.07 | 32.21 | 33.51 | 43 | No | negative |
| HP05596 | 20I/501Y.V1 | B.1.1.7 | 92.40 | 82.73 | 0.41 | 17.16 | 17.24 | 93 | Yes | negative |
| HP04615 | 20I/501Y.V1 | B.1.1.7 | 92.40 | 82.13 | 0.86 | 20.18 | 20.01 | 82 | yes | 3 |
| HP04999 | 20I/501Y.V1 | B.1.1.7 | 92.19 | 252.69 | not done | not done | not done | 27 | yes | negative |
| HP05494 | 20C | B.1.526.3 | 91.62 | 198.59 | 3.81 | 26.81 | 26.93 | 68 | Yes | negative |
| HP05012 | 20I/501Y.V1 | B.1.1.7 | 90.58 | 212.49 | 3.59 | 37.32 | 38.07 | 71 | yes | negative |

| | | | | | | | | | | |
|---|---|---|---|---|---|---|---|---|---|---|
| HP04298 | 20H/501Y.V2 | B.1.351 | 89.74 | 159.01 | not done | not done | not done | 85 | No | negative |
| HP04949 | 20I/501Y.V1 | B.1.1.7 | 88.91 | 198.58 | 4.18 | 30.13 | 30.11 | 9 | yes | negative |
| HP04970 | 20C | B.1.526.1 | 88.72 | 256.38 | 5.58 | 30.12 | 30.70 | 16 | yes | negative |
| HP04634 | 20I/501Y.V1 | B.1.1.7 | 88.65 | 73.38 | 1.00 | 22.37 | 22.67 | 75 | no | negative |
| HP04423 | 20I/501Y.V1 | B.1.1.7 | 87.94 | 194.91 | 6.52 | 26.96 | 26.77 | 28 | no | negative |
| HP02642 | 20B | B.1.1.207 | 86.39 | 178.70 | not done | not done | not done | 12 | no | negative |
| HP05620 | 20I/501Y.V1 | B.1.1.7 | 82.86 | 139.31 | not done | not done | not done | 31 | No | negative |
| HP04646 | 20A | B.1.153 | 80.62 | 52.47 | not done | not done | not done | 68 | yes | negative |
| HP05213 | 20I/501Y.V1 | B.1.1.7 | 79.58 | 152.19 | 1.19 | 31.91 | 31.93 | 49 | Yes | negative |
| HP04938 | 20I/501Y.V1 | B.1.1.7 | 77.30 | 146.08 | not done | not done | not done | 79 | no | negative |
| HP05100 | 20I/501Y.V1 | B.1.1.7 | 76.81 | 180.71 | 0.32 | 29.66 | 26.96 | 83 | No | negative |
| HP03628 | 20I/501Y.V1 | B.1.1.7 | 75.63 | 118.37 | 0.92 | 30.89 | 31.35 | 61 | yes | negative |
| HP05060 | 20J/501Y.V3 | B.1.566 | 75.58 | 151.32 | 0.50 | 29.04 | 28.58 | 3 | No | negative |
| HP04192 | 20G | B.1.2 | 70.05 | 90.64 | not done | not done | not done | 67 | No | negative |
| HP05061 | 20J/501Y.V3 | B.1.566 | 68.34 | 99.77 | 0.76 | 30.65 | 31.41 | 38 | No | negative |
| HP03047 | 20A | B.1.243 | 68.20 | 82.34 | 1.45 | 30.71 | 32.04 | 19 | no | negative |
| HP03839 | 20C | B.1 | 67.63 | 91.57 | 0.65 | 36.99 | 37.69 | 33 | no | negative |
| HP04636 | 20C | B.1 | 59.60 | 52.07 | 0.91 | 30.22 | 29.33 | 4 | yes | negative |
| HP04984 | 20I/501Y.V1 | B.1.1.7 | 56.74 | 113.05 | 0.94 | 30.82 | 28.09 | 68 | yes | negative |
| HP03826 | 20C | B.1 | 56.65 | 54.06 | 0.79 | 35.94 | ND | 75 | no | negative |
| HP04082 | 20A | B.1.165 | 55.79 | 51.65 | 1.48 | 34.85 | 35.71 | 69 | no | negative |
| HP04985 | 20I/501Y.V1 | B.1.1.7 | 51.33 | 62.29 | 2.49 | 32.46 | 31.88 | 66 | no | negative |
| HP02196 | 20C | None | 44.19 | 29.00 | 2.49 | 33.03 | 33.55 | 4 | yes | negative |
| HP05821 | 20A | None | 43.97 | 25.63 | 0.72 | 34.18 | 31.63 | 70 | yes | negative |
| HP04824 | 20I/501Y.V1 | None | 36.67 | 30.13 | 0.84 | 34.98 | 35.22 | 63 | no | negative |
| HP03590 | 20A | None | 31.41 | 23.80 | 4.05 | 34.25 | 34.66 | 30 | yes | negative |
| HP04598 | 20H/501Y.V2 | None | 26.20 | 13.67 | 2.51 | 27.31 | 27.55 | 49 | yes | negative |
| HP05059 | 20A | None | 25.99 | 18.95 | 4.79 | 35.98 | 34.63 | 2 | Yes | negative |
| HP05103 | 20A | None | 25.36 | 16.51 | 0.71 | 29.71 | 30.39 | 40 | No | negative |
| HP03529 | 20I/501Y.V1 | None | 23.64 | 16.11 | not done | not done | not done | 20 | no | negative |
| HP05191 | 20I/501Y.V1 | None | 22.33 | 20.69 | 1.17 | ND | ND | 56 | Yes | negative |
| HP03417 | 20C | None | 19.10 | 16.87 | 1.32 | 37.90 | 38.70 | 43 | yes | negative |
| HP05086 | 20I/501Y.V1 | None | 18.99 | 15.06 | 5.67 | ND | 37.29 | 21 | yes | negative |
| HP04629 | 20I/501Y.V1 | None | 15.72 | 9.56 | 1.74 | 36.45 | 35.76 | 50 | no | negative |

| | | | | | | | | | | |
|---|---|---|---|---|---|---|---|---|---|---|
| HP05808 | 20G | None | 15.60 | 9.99 | 6.32 | 31.44 | 30.51 | 33 | no | negative |
| HP04619 | 20B | None | 14.62 | 7.65 | 3.53 | 32.46 | 32.26 | 71 | no | negative |
| HP04939 | 20I/501Y.V1 | None | 12.69 | 10.04 | not done | not done | not done | 63 | no | negative |
| HP05010 | 20B | None | 12.68 | 8.46 | 1.35 | 37.41 | ND | 66 | no | negative |
| HP04835 | 20A | None | 9.16 | 11.43 | 5.06 | 37.59 | 36.32 | 9 | no | negative |
| HP02131 | 20A | None | 5.71 | 4.00 | not done | not done | not done | 14 | no | negative |
| HP02344 | 19A | None | 2.30 | 0.00 | not done | not done | not done | 31 | yes | negative |
| HP05613 | 20A | None | 2.21 | 1.51 | not done | not done | not done | 16 | Yes | negative |
| HP03057 | 20A | None | 1.11 | 1.65 | 5.41 | 32.72 | 32.14 | 20 | yes | negative |
| | Blue; potential false positives or very low viral loads | | | | | | | | | |
| HP03657 | 0 | 0 | 0.00 | 0.00 | 4.46 | ND | ND | 18 | no | negative |
| HP03664 | 0 | 0 | 0.00 | 0.00 | 4.59 | ND | ND | 30 | yes | negative |
| HP03671 | 0 | 0 | 0.00 | 0.00 | 1.02 | ND | ND | 54 | no | negative |
| HP04832 | 0 | 0 | 0.00 | 0.00 | 0.68 | 37.92 | 38.30 | 100 | yes | negative |
| HP05813 | 0 | 0 | 0.00 | 0.00 | 2.51 | ND | 38.32 | 37 | no | negative |
| HP05819 | 0 | 0 | 0.00 | 0.00 | 0.51 | 38.35 | 36.72 | 14 | no | negative |
| HP02185 | 0 | 0 | 0.00 | 0.00 | 5.23 | 36.22 | 38.01 | 5 | yes | negative |
| HP02170 | 0 | 0 | 0.00 | 0.00 | 5.50 | 38.06 | 36.32 | 14 | no | negative |
| HP02164 | 0 | 0 | 0.00 | 0.00 | 6.05 | 40.17 | 37.14 | 11 | yes | negative |
| HP02755 | 0 | None | 0.00 | 1.08 | 4.33 | 36.26 | 36.50 | 6 | yes | negative |
| HP03610 | 0 | 0 | 0.00 | 0.00 | 3.61 | ND | 38.55 | 9 | yes | negative |
| HP03831 | 0 | 0 | 0.00 | 0.00 | 3.21 | 34.43 | 34.91 | 41 | yes | negative |
| HP03849 | 0 | 0 | 0.00 | 0.00 | 1.08 | 36.90 | 37.70 | 66 | no | negative |
| HP04040 | 0 | 0 | 0.00 | 0.00 | 1.05 | ND | ND | 21 | no | negative |
| HP04235 | 0 | 0 | 0.00 | 0.00 | 4.52 | 38.14 | 38.83 | 75 | no | negative |
| HP04446 | 0 | 0 | 0.00 | 0.00 | 0.99 | ND | ND | 64 | no | negative |
| HP04451 | 0 | 0 | 0.00 | 0.00 | 1.23 | ND | ND | 37 | no | negative |
| HP04455 | 0 | 0 | 0.00 | 0.00 | 3.36 | 35.00 | 34.55 | 3 | no | negative |
| HP04614 | 0 | None | 0.00 | 0.14 | 3.04 | 35.46 | 34.32 | 6 | no | negative |
| HP05249 | 0 | 0 | 0.00 | 0.00 | not done | not done | not done | 4 | No | negative |
| HP05266 | 0 | None | 0.00 | 0.52 | 0.71 | 36.37 | 35.81 | 77 | No | negative |
| HP05484 | 0 | 0 | 0.00 | 0.00 | 0.30 | 32.35 | 32.34 | 4 | Yes | negative |
| HP04564 | 0 | 0 | 0.00 | 0.00 | not done | not done | not done | 40 | No | negative |
| HP02551 | 0 | 0 | 0.00 | 0.00 | 5.76 | ND | ND | 9 | no | negative |

**not done; no left over samples were available for serology or PerkinElmers PCR runs**
**ND; not detected**

Supplemental Table 2

| HPID | Clade | Pangolin_lineage_2_24 | %Coverage | Depth | IgG (OD) | PE ORF1ab Ct | PE N Ct | Symptoms | Days to CPE |
|---|---|---|---|---|---|---|---|---|---|
| HP02591 | 20G | B.1 | 98.60 | 350.17 | 0.78 | 16.66 | 16.70 | yes | 2 |
| HP02411 | 20I/501Y.V1 | B.1.1.7 | 99.60 | 443.00 | 0.49 | 23.46 | 21.37 | yes | 0 |
| HP02064 | 20I/501Y.V1 | B.1.1.7 | 99.60 | 389.00 | 3.77 | 24.79 | 23.98 | yes | 0 |
| HP02221 | 20I/501Y.V1 | B.1.1.7 | 99.60 | 445.00 | 0.33 | 19.84 | 18.96 | yes | 4 |
| HP02227 | 20I/501Y.V1 | B.1.1.7 | 99.60 | 437.00 | 0.32 | 21.12 | 20.35 | yes | 4 |
| HP02269 | 20I/501Y.V1 | B.1.1.7 | 99.60 | 402.00 | 0.48 | 21.55 | 18.90 | yes | 0 |
| HP02273 | 20I/501Y.V1 | B.1.1.7 | 99.60 | 314.00 | 0.41 | 22.01 | 21.01 | yes | 0 |
| HP02360 | 20I/501Y.V1 | B.1.1.7 | 99.60 | 447.00 | 0.53 | 18.06 | 16.99 | yes | 2 |
| HP02354 | 20I/501Y.V1 | B.1.1.7 | 99.60 | 447.00 | 0.40 | 19.65 | 18.00 | yes | 0 |
| HP02486 | 20I/501Y.V1 | B.1.1.7 | 99.59 | 435.00 | 0.28 | 30.52 | 31.08 | no | 0 |
| HP02492 | 20I/501Y.V1 | B.1.1.7 | 96.31 | 204.00 | 0.29 | 29.68 | 29.15 | yes | 0 |
| HP02503 | 20I/501Y.V1 | B.1.1.7 | 97.31 | 420.00 | 0.58 | 26.91 | 25.85 | yes | 0 |
| HP02500 | 20I/501Y.V1 | B.1.1.7 | 99.60 | 443.00 | 0.44 | 16.92 | 14.63 | yes | 2 |
| HP02497 | 20I/501Y.V1 | B.1.1.7 | 99.60 | 435.00 | 0.39 | 16.51 | 15.46 | yes | 3 |
| HP02528 | 20I/501Y.V1 | B.1.1.7 | 97.31 | 261.00 | 0.23 | 26.51 | 24.98 | yes | 0 |
| HP02652 | 20I/501Y.V1 | B.1.1.7 | 99.60 | 442.44 | 0.38 | 23.26 | 22.70 | yes | 2 |
| HP02664 | 20I/501Y.V1 | B.1.1.7 | 99.60 | 399.73 | 0.40 | 28.43 | 26.73 | yes | 0 |
| HP02675 | 20I/501Y.V1 | B.1.1.7 | 92.55 | 236.07 | 0.40 | 31.50 | 31.13 | no | 0 |
| HP02714 | 20I/501Y.V1 | B.1.1.7 | 99.60 | 421.23 | 0.29 | 26.43 | 24.88 | yes | 0 |
| HP02639 | 20I/501Y.V1 | B.1.1.7 | 99.60 | 444.83 | 0.36 | 18.99 | 18.46 | yes | 5 |
| HP02638 | 20I/501Y.V1 | B.1.1.7 | 99.60 | 443.07 | 0.29 | 17.71 | 16.88 | yes | 3 |
| HP02792 | 20I/501Y.V1 | B.1.1.7 | 96.05 | 182.15 | 0.47 | 25.80 | 24.76 | yes | 0 |
| HP02689 | 20I/501Y.V1 | B.1.1.7 | 99.60 | 422.63 | 0.42 | 26.54 | 25.83 | yes | 0 |
| HP02724 | 20I/501Y.V1 | B.1.1.7 | 98.60 | 281.09 | 0.47 | 14.32 | 13.58 | yes | 3 |
| HP02723 | 20I/501Y.V1 | B.1.1.7 | 96.91 | 120.33 | 0.36 | 17.05 | 16.33 | yes | 2 |
| HP02727 | 20I/501Y.V1 | B.1.1.7 | 96.76 | 130.44 | 0.27 | 16.22 | 15.48 | yes | 2 |
| HP02725 | 20I/501Y.V1 | B.1.1.7 | 97.04 | 180.66 | 0.36 | 21.01 | 20.60 | yes | 4 |
| HP02767 | 20I/501Y.V1 | B.1.1.7 | 98.60 | 318.32 | 0.39 | 19.99 | 19.32 | yes | 3 |
| HP03027 | 20I/501Y.V1 | B.1.1.7 | 97.15 | 381.50 | 0.31 | 23.44 | 22.47 | yes | 3 |
| HP03075 | 20I/501Y.V1 | B.1.1.7 | 98.60 | 367.13 | 0.39 | 23.91 | 23.68 | yes | 0 |
| HP03077 | 20I/501Y.V1 | B.1.1.7 | 98.61 | 367.82 | 0.80 | 22.83 | 21.93 | yes | 0 |

| | | | | | | | | | |
|---|---|---|---|---|---|---|---|---|---|
| HP03134 | 20I/501Y.V1 | B.1.1.7 | 99.60 | 446.53 | 0.43 | 14.24 | 12.47 | yes | 3 |
| HP03135 | 20I/501Y.V1 | B.1.1.7 | 99.60 | 444.21 | 0.34 | 20.92 | 20.24 | yes | 4 |
| HP03026 | 20I/501Y.V1 | B.1.1.7 | 93.40 | 333.15 | 0.38 | 25.40 | 24.38 | yes | 0 |
| HP03132 | 20I/501Y.V1 | B.1.1.7 | 99.60 | 446.02 | 0.31 | 21.92 | 21.51 | yes | 4 |
| HP03131 | 20I/501Y.V1 | B.1.1.7 | 99.60 | 444.78 | 0.38 | 22.78 | 21.85 | yes | 0 |
| HP03180 | 20I/501Y.V1 | B.1.1.7 | 99.60 | 448.44 | 0.42 | 15.20 | 13.33 | no | 2 |
| HP03204 | 20I/501Y.V1 | B.1.1.7 | 99.60 | 440.39 | 0.36 | 24.53 | 24.09 | yes | 5 |
| HP03202 | 20I/501Y.V1 | B.1.1.7 | 99.60 | 440.13 | 0.41 | 24.51 | 22.85 | yes | 0 |
| HP03209 | 20I/501Y.V1 | B.1.1.7 | 99.60 | 447.93 | 0.44 | 19.00 | 18.15 | yes | 3 |
| HP03242 | 20I/501Y.V1 | B.1.1.7 | 99.59 | 445.94 | 0.37 | 15.19 | 14.14 | yes | 3 |
| HP03229 | 20I/501Y.V1 | B.1.1.7 | 99.60 | 438.28 | 0.31 | 24.34 | 23.27 | yes | 0 |
| HP03227 | 20I/501Y.V1 | B.1.1.7 | 98.71 | 414.71 | 0.33 | 29.20 | 27.65 | yes | 0 |
| HP03186 | 20I/501Y.V1 | B.1.1.7 | 99.59 | 449.38 | 0.47 | 16.48 | 15.46 | yes | 2 |
| HP03200 | 20I/501Y.V1 | B.1.1.7 | 99.60 | 446.60 | 0.30 | 21.79 | 20.50 | yes | 4 |
| HP03250 | 20I/501Y.V1 | B.1.1.7 | 99.60 | 447.18 | 0.36 | 18.67 | 17.72 | yes | 3 |
| HP02338 | 20G | B.1.2 | 99.60 | 428.00 | 0.35 | 16.86 | 16.37 | yes | 2 |
| HP02336 | 20G | B.1.2 | 99.60 | 410.00 | 0.25 | 15.94 | 15.91 | yes | 2 |
| HP02374 | 20G | B.1.2 | 99.60 | 449.00 | 0.36 | 16.93 | 16.49 | yes | 2 |
| HP02369 | 20G | B.1.2 | 99.60 | 446.00 | 0.26 | 21.20 | 18.80 | yes | 2 |
| HP02391 | 20G | B.1.2 | 99.60 | 449.00 | 0.27 | 12.79 | 12.61 | yes | 2 |
| HP02372 | 20G | B.1.2 | 98.87 | 444.00 | 0.21 | 17.24 | 16.14 | yes | 2 |
| HP02367 | 20G | B.1.2 | 99.02 | 397.00 | 0.31 | 26.09 | 25.73 | yes | 0 |
| HP02388 | 20G | B.1.2 | 99.60 | 448.00 | 0.34 | 17.55 | 15.75 | yes | 2 |
| HP02390 | 20G | B.1.2 | 99.60 | 440.00 | 0.31 | 23.96 | 23.53 | yes | 2 |
| HP02386 | 20G | B.1.2 | 99.60 | 449.00 | 0.28 | 15.41 | 15.11 | no | 2 |
| HP02407 | 20G | B.1.2 | 99.59 | 447.00 | 0.37 | 17.65 | 17.01 | yes | 2 |
| HP02412 | 20G | B.1.2 | 99.60 | 439.00 | 0.22 | 24.66 | 24.19 | no | 3 |
| HP02404 | 20G | B.1.2 | 99.60 | 446.00 | 1.16 | 13.79 | 13.05 | yes | 2 |
| HP02469 | 20G | B.1.2 | 99.60 | 443.00 | 0.31 | 19.68 | 19.46 | yes | 2 |
| HP02397 | 20G | B.1.2 | 99.60 | 444.00 | 0.35 | 23.28 | 23.17 | no | 3 |
| HP02389 | 20G | B.1.2 | 97.87 | 324.00 | 0.35 | 28.15 | 27.32 | yes | 0 |
| HP02525 | 20G | B.1.2 | 91.74 | 141.00 | 1.89 | 30.59 | 30.03 | no | 0 |
| HP02459 | 20G | B.1.2 | 99.60 | 420.00 | 0.34 | 21.81 | 22.00 | yes | 3 |
| HP02517 | 20G | B.1.2 | 99.59 | 352.00 | 0.31 | 20.70 | 20.43 | yes | 2 |

| | | | | | | | | | |
|---|---|---|---|---|---|---|---|---|---|
| HP02443 | 20G | B.1.2 | 94.36 | 228.00 | 0.26 | 31.21 | 31.02 | yes | 0 |
| HP02477 | 20G | B.1.2 | 98.60 | 308.00 | 0.35 | 26.76 | 26.75 | yes | 0 |
| HP02347 | 20G | B.1.2 | 99.60 | 445.00 | 0.36 | 20.05 | 19.57 | yes | 2 |
| HP02355 | 20G | B.1.2 | 99.60 | 445.00 | 0.27 | 17.03 | 16.33 | yes | 2 |
| HP02489 | 20G | B.1.2 | 95.91 | 225.00 | 1.79 | 29.29 | 29.33 | no | 0 |
| HP02356 | 20G | B.1.2 | 99.60 | 446.00 | 0.34 | 15.97 | 15.99 | no | 2 |
| HP02555 | 20G | B.1.2 | 95.98 | 236.24 | 0.20 | 21.18 | 19.70 | yes | 2 |
| HP02501 | 20G | B.1.2 | 97.31 | 394.00 | 0.27 | 24.95 | 24.75 | yes | 2 |
| HP02544 | 20G | B.1.2 | 91.62 | 208.79 | 0.57 | 28.46 | 28.76 | no | 0 |
| HP02363 | 20G | B.1.2 | 99.60 | 448.00 | 0.73 | 16.94 | 16.01 | yes | 4 |
| HP02364 | 20G | B.1.2 | 99.60 | 445.00 | 0.29 | 20.85 | 20.68 | yes | 2 |
| HP02506 | 20G | B.1.2 | 94.57 | 246.00 | 0.20 | 31.73 | 31.34 | yes | 0 |
| HP02548 | 20G | B.1.2 | 97.61 | 301.74 | 0.53 | 23.47 | 22.82 | yes | 0 |
| HP02509 | 20G | B.1.2 | 98.68 | 425.00 | 0.39 | 20.24 | 19.97 | yes | 3 |
| HP02570 | 20G | B.1.2 | 97.61 | 278.82 | 0.81 | 29.71 | 29.30 | yes | 0 |
| HP02571 | 20G | B.1.2 | 97.61 | 315.33 | 0.21 | 24.70 | 23.95 | no | 0 |
| HP02533 | 20G | B.1.2 | 97.61 | 270.55 | 0.22 | 16.12 | 16.15 | yes | 2 |
| HP02536 | 20G | B.1.2 | 98.60 | 312.55 | 0.27 | 18.25 | 18.17 | yes | 2 |
| HP02535 | 20G | B.1.2 | 99.60 | 350.42 | 0.43 | 15.55 | 14.86 | no | 2 |
| HP02585 | 20G | B.1.2 | 94.54 | 178.95 | 0.22 | 29.58 | 27.15 | yes | 0 |
| HP02577 | 20G | B.1.2 | 97.61 | 307.10 | 0.25 | 25.41 | 25.04 | no | 3 |
| HP02607 | 20G | B.1.2 | 96.82 | 297.60 | 0.67 | 22.29 | 21.76 | yes | 0 |
| HP02589 | 20G | B.1.2 | 99.60 | 387.50 | 0.62 | 20.16 | 19.76 | yes | 2 |
| HP02653 | 20G | B.1.2 | 99.60 | 416.37 | 0.41 | 26.80 | 26.77 | yes | 0 |
| HP02657 | 20G | B.1.2 | 94.03 | 262.27 | 5.11 | 32.38 | 29.07 | yes | 0 |
| HP02710 | 20G | B.1.2 | 99.60 | 426.28 | 0.27 | 19.87 | 19.05 | yes | 3 |
| HP02651 | 20G | B.1.2 | 99.60 | 424.19 | 0.37 | 24.43 | 24.25 | yes | 3 |
| HP02628 | 20G | B.1.2 | 98.61 | 439.84 | 0.26 | 15.53 | 15.23 | yes | 2 |
| HP02682 | 20G | B.1.2 | 99.60 | 432.70 | 1.06 | 24.52 | 24.71 | yes | 0 |
| HP02698 | 20G | B.1.2 | 96.99 | 261.86 | 0.50 | 26.26 | 26.22 | yes | 0 |
| HP02685 | 20G | B.1.2 | 99.60 | 409.73 | 0.35 | 23.71 | 23.88 | yes | 0 |
| HP02683 | 20G | B.1.2 | 99.60 | 436.39 | 3.32 | 22.67 | 22.86 | yes | 5 |
| HP02604 | 20G | B.1.2 | 97.61 | 321.34 | 0.29 | 20.14 | 20.03 | yes | 2 |
| HP02713 | 20G | B.1.2 | 99.60 | 447.87 | 0.35 | 15.86 | 15.49 | yes | 2 |

| | | | | | | | | | |
|---|---|---|---|---|---|---|---|---|---|
| HP02635 | 20G | B.1.2 | 99.60 | 443.34 | 0.58 | 16.69 | 16.29 | yes | 3 |
| HP02735 | 20G | B.1.2 | 97.99 | 228.56 | 0.50 | 17.00 | 16.27 | yes | 2 |
| HP02765 | 20G | B.1.2 | 98.66 | 370.15 | 0.30 | 21.49 | 21.52 | yes | 4 |
| HP02762 | 20G | B.1.2 | 98.03 | 336.60 | 0.29 | 23.20 | 23.98 | yes | 3 |
| HP02751 | 20G | B.1.2 | 97.68 | 345.15 | 0.51 | 28.07 | 29.02 | yes | 4 |
| HP02721 | 20G | B.1.2 | 99.60 | 397.69 | 0.58 | 15.85 | 15.80 | yes | 2 |
| HP02775 | 20G | B.1.2 | 97.85 | 186.10 | 0.30 | 25.92 | 25.69 | yes | 0 |
| HP02772 | 20G | B.1.2 | 99.59 | 413.38 | 0.49 | 18.71 | 18.37 | yes | 2 |
| HP02776 | 20G | B.1.2 | 98.60 | 295.23 | 0.35 | 20.71 | 20.29 | yes | 2 |
| HP03031 | 20G | B.1.2 | 93.45 | 365.22 | 0.36 | 23.63 | 23.87 | yes | 2 |
| HP03067 | 20G | B.1.2 | 98.41 | 374.71 | 0.24 | 25.35 | 25.35 | yes | 3 |
| HP03060 | 20G | B.1.2 | 96.62 | 222.17 | 0.35 | 21.70 | 21.24 | yes | 3 |
| HP03036 | 20G | B.1.2 | 99.60 | 422.96 | 0.44 | 16.42 | 16.62 | no | 2 |
| HP03103 | 20G | B.1.2 | 99.60 | 442.02 | 0.41 | 16.94 | 16.73 | yes | 2 |
| HP03113 | 20G | B.1.2 | 98.60 | 344.66 | 0.49 | 28.00 | 28.74 | yes | 0 |
| HP03109 | 20G | B.1.2 | 99.60 | 445.28 | 0.92 | 27.83 | 27.90 | yes | 0 |
| HP03107 | 20G | B.1.2 | 99.60 | 444.55 | 0.42 | 14.85 | 14.83 | yes | 2 |
| HP03148 | 20G | B.1.2 | 99.02 | 374.78 | 0.35 | 24.61 | 25.44 | yes | 3 |
| HP03122 | 20G | B.1.2 | 99.60 | 443.96 | 0.27 | 20.39 | 20.96 | yes | 3 |
| HP03130 | 20G | B.1.2 | 98.09 | 335.66 | 0.87 | 28.89 | 27.98 | yes | 0 |
| HP03212 | 20G | B.1.2 | 99.60 | 444.50 | 1.40 | 19.87 | 20.03 | No | 2 |
| HP03215 | 20G | B.1.2 | 99.44 | 441.19 | 0.89 | 18.46 | 18.44 | yes | 2 |
| HP03086 | 20H/501Y.V2 | B.1.351 | 95.89 | 191.56 | 0.38 | 23.89 | 24.17 | yes | 4 |
| HP03124 | 20H/501Y.V2 | B.1.351 | 98.87 | 388.28 | 0.37 | 27.27 | 27.23 | yes | 0 |
| HP03201 | 20H/501Y.V2 | B.1.351 | 98.87 | 442.12 | 0.36 | 23.73 | 24.44 | yes | 3 |

Supplemental Table 3

| HPID | EPI_ISL | Lineage | substitutic | AA deletions |
|---|---|---|---|---|
| **Fully Vaccinated** | | | | |
| HP05084 | EPI_ISL_19 | B.1.526.1 | N:M1X,N:T | ORF1a:S3675-,ORF1a:G3676-,ORF1a:F3677- |
| HP05581 | EPI_ISL_21 | B.1.526 | N:M1X,N:P | ORF1a:S3675-,ORF1a:G3676-,ORF1a:F3677- |
| HP05624 | | B.1.2 | N:M1X,N:P | ORF1a:S3675-,ORF1a:G3676-,ORF1a:F3677- |
| HP04923 | EPI_ISL_18 | B.1.2 | N:P67S,N:P199L,N:D377Y,ORF1a:T265I,ORF1a:T1246I,ORF1a:M2606I,ORF1a:L3352F,ORF1b:P314L,ORF1b:T116 | |
| HP04957 | | B.1.1.7 | N:M1X,N:D | ORF1a:S3675-,ORF1a:G3676-,ORF1a:F3677-,S:H69-,S:V70-,S:Y144- |
| HP04198 | EPI_ISL_16 | B.1.596 | E:L21V,N:P67S,N:P199L,N:T271I,N:E378Q,ORF1a:T265I,ORF1a:E974G,ORF1a:T2280I,ORF1a:L3352F,ORF1b:P314 | |
| HP05012 | EPI_ISL_19 | B.1.1.7 | N:M1X,N:D | ORF1a:S3675-,ORF1a:G3676-,ORF1a:F3677-,S:H69-,S:V70-,S:Y144- |
| HP05105 | EPI_ISL_19 | B.1.1.7 | N:M1X,N:D | ORF1a:S3675-,ORF1a:G3676-,ORF1a:F3677-,S:H69-,S:V70-,S:Y144- |
| HP05805 | | B.1.1.7 | N:M1X,N:D | ORF1a:S3675-,ORF1a:G3676-,ORF1a:F3677-,S:H69-,S:V70-,S:Y144- |
| HP04818 | EPI_ISL_18 | B.1.1.7 | N:M1X,N:D | ORF1a:S3675-,ORF1a:G3676-,ORF1a:F3677-,S:H69-,S:V70-,S:Y144- |
| HP05815 | EPI_ISL_22 | B.1.1.7 | N:M1X,N:D | ORF1a:S3675-,ORF1a:G3676-,ORF1a:F3677-,S:H69-,S:V70-,S:Y144- |
| HP05816 | EPI_ISL_22 | B.1.526 | N:M1X,N:P | ORF1a:S3675-,ORF1a:G3676-,ORF1a:F3677- |
| HP03729 | EPI_ISL_14 | B.1.1.7 | N:M1X,N:D | ORF1a:S3675-,ORF1a:G3676-,ORF1a:F3677-,S:H69-,S:V70-,S:Y144- |
| HP03775 | EPI_ISL_14 | B.1.1.7 | N:M1X,N:D | ORF1a:S3675-,ORF1a:G3676-,ORF1a:F3677-,S:H69-,S:V70-,S:Y144- |
| HP03786 | EPI_ISL_14 | B.1.1.7 | N:M1X,N:D | ORF1a:S3675-,ORF1a:G3676-,ORF1a:F3677-,S:H69-,S:V70-,S:Y144- |
| HP04129 | EPI_ISL_15 | B.1.1.7 | N:M1X,N:D | ORF1a:S3675-,ORF1a:G3676-,ORF1a:F3677-,S:H69-,S:V70-,S:Y144- |
| HP04153 | EPI_ISL_19 | B.1.1.7 | M:G89S,N: | ORF1a:S3675-,ORF1a:G3676-,ORF1a:F3677-,S:H69-,S:V70-,S:Y144- |
| HP04182 | EPI_ISL_19 | B.1.1.7 | N:M1X,N:D | ORF1a:S3675-,ORF1a:G3676-,ORF1a:F3677-,S:H69-,S:V70-,S:Y144- |
| HP04113 | EPI_ISL_15 | B.1.526 | N:M1X,N:P | ORF1a:S3675-,ORF1a:G3676-,ORF1a:F3677- |
| HP04414 | EPI_ISL_16 | B.1.1.7 | N:M1X,N:D | ORF1a:S3675-,ORF1a:G3676-,ORF1a:F3677-,S:H69-,S:V70-,S:Y144- |
| HP04499 | EPI_ISL_16 | B.1.1.7 | N:M1X,N:D | ORF1a:S3675-,ORF1a:G3676-,ORF1a:F3677-,S:H69-,S:V70-,S:Y144- |
| HP04930 | EPI_ISL_18 | B.1.1.7 | N:M1X,N:D | ORF1a:S3675-,ORF1a:G3676-,ORF1a:F3677-,S:H69-,S:V70-,S:Y144- |
| HP04931 | EPI_ISL_18 | B.1.1.7 | N:M1X,N:D | ORF1a:S3675-,ORF1a:G3676-,ORF1a:F3677-,S:H69-,S:V70-,S:Y144- |
| HP05000 | EPI_ISL_19 | B.1.526 | N:M1X,N:P | ORF1a:S3675-,ORF1a:G3676-,ORF1a:F3677- |
| HP05016 | | B.1.1.7 | N:M1X,N:D | ORF1a:S3675-,ORF1a:G3676-,ORF1a:F3677-,S:H69-,S:V70-,S:Y144- |
| HP05024 | EPI_ISL_19 | B.1.1.7 | N:M1X,N:D | ORF1a:S3675-,ORF1a:G3676-,ORF1a:F3677-,S:H69-,S:V70-,S:Y144- |
| HP04481 | EPI_ISL_16 | P.1 | N:P80R,N:I | ORF1a:S3675-,ORF1a:G3676-,ORF1a:F3677- |
| HP05031 | EPI_ISL_19 | B.1.1.7 | N:M1X,N:D | ORF1a:S3675-,ORF1a:G3676-,ORF1a:F3677-,S:H69-,S:V70-,S:Y144- |
| HP03722 | EPI_ISL_19 | B.1.526 | N:M1X,N:P | ORF1a:S3675-,ORF1a:G3676-,ORF1a:F3677- |
| HP04993 | EPI_ISL_21 | B.1.214.2 | N:M1X,N:D | ORF1a:S3675-,ORF1a:G3676-,ORF1a:F3677- |
| HP05512 | EPI_ISL_21 | B.1.1.7 | N:M1X,N:D | ORF1a:S3675-,ORF1a:G3676-,ORF1a:F3677-,S:H69-,S:V70-,S:Y144- |

| | | | | |
|---|---|---|---|---|
| HP05025 | EPI_ISL_19 | B.1.620 | N:A220V,O | ORF1a:S3675-,ORF1a:G3676-,ORF1a:F3677-,ORF7b:L14-,S:H69-,S:V70-,S:Y144-,S:L242-,S:A243- |
| HP03453 | EPI_ISL_14 | B.1.1.7 | N:M1X,N:D | ORF1a:S3675-,ORF1a:G3676-,ORF1a:F3677-,S:H69-,S:V70-,S:Y144- |
| HP04204 | EPI_ISL_16 | B.1.1.7 | N:M1X,N:D | ORF1a:S3675-,ORF1a:G3676-,ORF1a:F3677-,S:H69-,S:V70-,S:Y144- |
| HP04320 | EPI_ISL_16 | B.1.1.7 | N:M1X,N:D | ORF1a:S3675-,ORF1a:G3676-,ORF1a:F3677-,S:H69-,S:V70-,S:Y144- |
| HP04461 | EPI_ISL_16 | B.1.1.7 | N:M1X,N:D | ORF1a:S3675-,ORF1a:G3676-,ORF1a:F3677-,S:H69-,S:V70-,S:Y144- |
| HP04464 | | B.1.1.7 | N:M1X,N:D | ORF1a:S3675-,ORF1a:G3676-,ORF1a:F3677-,S:H69-,S:V70-,S:Y144- |
| HP05130 | EPI_ISL_19 | B.1.1.7 | N:M1X,N:D | ORF1a:S3675-,ORF1a:G3676-,ORF1a:F3677-,S:H69-,S:V70-,S:Y144- |
| HP05159 | EPI_ISL_19 | B.1.1.7 | N:M1X,N:D | ORF1a:S3675-,ORF1a:G3676-,ORF1a:F3677-,S:H69-,S:V70-,S:Y144- |
| HP05181 | EPI_ISL_19 | B.1.1.7 | N:M1X,N:D | ORF1a:S3675-,ORF1a:G3676-,ORF1a:F3677-,S:H69-,S:V70-,S:Y144- |
| HP05595 | EPI_ISL_21 | B.1.1.7 | N:M1X,N:D | ORF1a:S3675-,ORF1a:G3676-,ORF1a:F3677-,S:H69-,S:V70-,S:Y144- |
| HP04826 | EPI_ISL_18 | B.1.1.7 | N:M1X,N:D | ORF1a:S3675-,ORF1a:G3676-,ORF1a:F3677-,S:H69-,S:V70-,S:Y144- |
| HP05129 | EPI_ISL_19 | B.1.526 | N:M1X,N:P | ORF1a:S3675-,ORF1a:G3676-,ORF1a:F3677- |
| HP04656 | EPI_ISL_23 | B.1.1.7 | N:M1X,N:D | ORF1a:S3675-,ORF1a:G3676-,ORF1a:F3677-,S:H69-,S:V70-,S:Y144- |
| HP04632 | EPI_ISL_23 | B.1.1.7 | N:M1X,N:D | ORF1a:S3675-,ORF1a:G3676-,ORF1a:F3677-,S:H69-,S:V70-,S:Y144- |
| HP05509 | EPI_ISL_21 | B.1.351 | E:P71L,N:P | ORF1a:S3675-,ORF1a:G3676-,ORF1a:F3677-,S:A243-,S:L244-,S:H245- |
| HP05160 | EPI_ISL_19 | B.1.1.7 | N:M1X,N:D | ORF1a:S3675-,ORF1a:G3676-,ORF1a:F3677-,S:H69-,S:V70-,S:Y144- |
| HP05201 | EPI_ISL_19 | B.1.1.7 | N:M1X,N:D | ORF1a:S3675-,ORF1a:G3676-,ORF1a:F3677-,ORF7a:T120-,ORF7a:E121-,ORF7a:*122-,ORF7b:M1-,OR |
| HP04665 | EPI_ISL_16 | B.1.526.1 | N:M1X,N:T | ORF1a:S3675-,ORF1a:G3676-,ORF1a:F3677- |
| HP04430 | EPI_ISL_19 | B.1.1.7 | N:M1X,N:D | ORF1a:S3675-,ORF1a:G3676-,ORF1a:F3677-,ORF7a:S60-,ORF7a:T61-,ORF7a:Q62-,ORF7a:F63-,ORF7a |
| HP05168 | EPI_ISL_19 | B.1.1.7 | N:M1X,N:D | ORF1a:S3675-,ORF1a:G3676-,ORF1a:F3677-,S:H69-,S:V70-,S:Y144- |
| HP05171 | EPI_ISL_19 | B.1.1.7 | N:M1X,N:D | ORF1a:S3675-,ORF1a:G3676-,ORF1a:F3677-,S:H69-,S:V70-,S:Y144- |
| HP05145 | EPI_ISL_19 | P.1 | N:P80R,N:I | ORF1a:S3675-,ORF1a:G3676-,ORF1a:F3677- |
| HP03053 | EPI_ISL_12 | B.1.1.7 | N:M1X,N:D | ORF1a:S3675-,ORF1a:G3676-,ORF1a:F3677-,S:H69-,S:V70-,S:Y144- |
| HP05277 | EPI_ISL_21 | B.1.526 | N:M1X,N:P | ORF1a:S3675-,ORF1a:G3676-,ORF1a:F3677- |
| HP04074 | EPI_ISL_15 | B.1.1.7 | N:M1X,N:D | ORF1a:S3675-,ORF1a:G3676-,ORF1a:F3677-,S:H69-,S:V70-,S:Y144- |
| HP02169 | EPI_ISL_14 | B.1.1 | N:R203K,N | ORF3a:V256- |
| HP03374 | EPI_ISL_13 | B.1.526.1 | N:M1X,N:P199L,N:M234I,ORF1a:T265I,ORF1a:L3201P,ORF1b:P314L,ORF3a:P42L,ORF3a:Q57H,ORF8:T11I,S:L5F, | |
| HP05596 | | B.1.1.7 | N:M1X,N:D | ORF1a:S3675-,ORF1a:G3676-,ORF1a:F3677-,S:H69-,S:V70-,S:Y144- |
| HP04615 | | B.1.1.7 | N:M1X,N:D | ORF1a:S3675-,ORF1a:G3676-,ORF1a:F3677-,S:H69-,S:V70-,S:Y144- |
| HP04999 | EPI_ISL_19 | B.1.1.7 | N:M1X,N:D | ORF1a:S3675-,ORF1a:G3676-,ORF1a:F3677-,S:H69-,S:V70-,S:Y144- |
| HP05494 | EPI_ISL_21 | B.1.526.3 | N:M1X,N:P | ORF1a:S3675-,ORF1a:G3676-,ORF1a:F3677- |
| HP04298 | | B.1.351 | E:P71L,N:T | ORF1a:S3675-,ORF1a:G3676-,ORF1a:F3677-,S:A243-,S:L244-,S:H245- |
| HP05604 | EPI_ISL_22 | B.1.526 | N:M1X,N:P | ORF1a:S3675-,ORF1a:G3676-,ORF1a:F3677- |
| HP05616 | EPI_ISL_22 | B.1.1.7 | N:M1X,N:D | ORF1a:S3675-,ORF1a:G3676-,ORF1a:F3677-,S:H69-,S:V70-,S:Y144- |

| | | | | |
|---|---|---|---|---|
| HP05279 | EPI_ISL_22 | B.1.1.7 | N:M1X,N:D | ORF1a:S3675-,ORF1a:G3676-,ORF1a:F3677-,S:H69-,S:V70-,S:Y144- |
| HP05253 | EPI_ISL_22 | B.1.575 | M:I82T,N:M1X,ORF1a:T265I,ORF1a:K367R,ORF1a:A687D,ORF1a:V2061I,ORF1a:K2395R,ORF1a:T3255I,ORF1a:T3 | |
| Control Group | | | | |
| HP03082 | EPI_ISL_12 | B.1.1.7 | N:M1X,N:D | ORF1a:S3675-,ORF1a:G3676-,ORF1a:F3677-,S:H69-,S:V70-,S:Y144- |
| HP03086 | EPI_ISL_12 | B.1.351 | E:P71L,N:T | ORF1a:S3675-,ORF1a:G3676-,ORF1a:F3677-,S:A243-,S:L244-,S:H245- |
| HP03247 | EPI_ISL_12 | B.1.429 | N:T205I,N:M234I,ORF1a:H110Y,ORF1a:T265I,ORF1a:I4205V,ORF1b:P314L,ORF1b:D1183Y,ORF3a:Q57H,ORF8:S6 | |
| HP03405 | EPI_ISL_13 | B.1.2 | N:P67S,N:P199L,ORF1a:T265I,ORF1a:L2295F,ORF1a:M2606I,ORF1a:E2918G,ORF1a:L3352F,ORF1a:M3752I,ORF1 | |
| HP03406 | EPI_ISL_13 | B.1.1.486 | N:R203K,N:G204R,N:M234I,ORF1a:S2822F,ORF1b:P314L,ORF3a:S58N,ORF3a:H78Y,ORF3a:P104L,ORF6:I14T,S:T | |
| HP03407 | EPI_ISL_13 | B.1.311 | N:Q9H,N:T135I,N:P207L,N:T391I,ORF1a:F143L,ORF1a:T265I,ORF1a:Q1582H,ORF1a:T2978I,ORF1a:T4087I,ORF1a | |
| HP03409 | EPI_ISL_13 | B.1.2 | N:P67S,N:P199L,N:P383L,ORF1a:T265I,ORF1a:L3352F,ORF1a:D3526G,ORF1b:P314L,ORF1b:N1653D,ORF1b:R21 | |
| HP03436 | EPI_ISL_13 | B.1.526.1 | N:M1X,N:T | ORF1a:S3675-,ORF1a:G3676-,ORF1a:F3677-,S:Y144- |
| HP03438 | EPI_ISL_13 | B.1.1.7 | N:M1X,N:D | ORF1a:S3675-,ORF1a:G3676-,ORF1a:F3677-,S:H69-,S:V70-,S:Y144- |
| HP03439 | EPI_ISL_13 | B.1.1.7 | N:M1X,N:D | ORF1a:S3675-,ORF1a:G3676-,ORF1a:F3677-,S:H69-,S:V70-,S:Y144- |
| HP03449 | EPI_ISL_14 | B.1.1.7 | N:M1X,N:D | ORF1a:S3675-,ORF1a:G3676-,ORF1a:F3677-,S:H69-,S:V70-,S:Y144- |
| HP03450 | EPI_ISL_14 | B.1.243 | N:S194L,ORF1a:V438A,ORF1a:G2118D,ORF1a:N2523T,ORF1b:P314L,ORF1b:A576V,ORF3a:T151I,ORF6:K42N,S:D | |
| HP03530 | EPI_ISL_14 | B.1.351 | E:P71L,N:T | ORF1a:S3675-,ORF1a:G3676-,ORF1a:F3677-,S:A243-,S:L244-,S:H245- |
| HP03840 | EPI_ISL_19 | B.1.526.1 | N:M1X,N:P | ORF1a:S3675-,ORF1a:G3676-,ORF1a:F3677- |
| HP03862 | EPI_ISL_14 | B.1.1.7 | N:M1X,N:D | ORF1a:S3675-,ORF1a:G3676-,ORF1a:F3677-,S:H69-,S:V70-,S:Y144- |
| HP03875 | EPI_ISL_14 | B.1.526.1 | N:M1X,N:T | ORF1a:S3675-,ORF1a:G3676-,ORF1a:F3677- |
| HP03877 | EPI_ISL_14 | B.1.526.1 | N:M1X,N:T | ORF1a:S3675-,ORF1a:G3676-,ORF1a:F3677- |
| HP03879 | EPI_ISL_14 | B.1.526.1 | N:M1X,N:T | ORF1a:S3675-,ORF1a:G3676-,ORF1a:F3677- |
| HP04004 | EPI_ISL_15 | B.1.1.7 | N:M1X,N:D | ORF1a:S3675-,ORF1a:G3676-,ORF1a:F3677-,S:H69-,S:V70-,S:Y144- |
| HP04005 | EPI_ISL_15 | B.1.1.7 | N:M1X,N:D | ORF1a:S3675-,ORF1a:G3676-,ORF1a:F3677-,S:H69-,S:V70-,S:Y144- |
| HP04051 | EPI_ISL_15 | B.1.1.7 | N:M1X,N:D | ORF1a:S3675-,ORF1a:G3676-,ORF1a:F3677-,S:H69-,S:V70-,S:Y144- |
| HP04053 | EPI_ISL_15 | B.1.526.1 | N:M1X,N:T | ORF1a:S3675-,ORF1a:G3676-,ORF1a:F3677- |
| HP04054 | EPI_ISL_15 | B.1.1.7 | N:M1X,N:D | ORF1a:S3675-,ORF1a:G3676-,ORF1a:F3677-,S:H69-,S:V70-,S:Y144- |
| HP04056 | EPI_ISL_15 | B.1.1.7 | N:M1X,N:D | ORF1a:S3675-,ORF1a:G3676-,ORF1a:F3677-,S:H69-,S:V70-,S:Y144- |
| HP04057 | EPI_ISL_15 | B.1.1.7 | N:M1X,N:D | ORF1a:S3675-,ORF1a:G3676-,ORF1a:F3677-,S:H69-,S:V70-,S:Y144- |
| HP04062 | EPI_ISL_15 | B.1.526.1 | N:M1X,N:P | ORF1a:S3675-,ORF1a:G3676-,ORF1a:F3677- |
| HP04063 | EPI_ISL_15 | B.1.1.7 | N:M1X,N:D | ORF1a:S3675-,ORF1a:G3676-,ORF1a:F3677-,S:H69-,S:V70-,S:Y144- |
| HP04064 | EPI_ISL_15 | B.1.1.7 | N:M1X,N:D | ORF1a:S3675-,ORF1a:G3676-,ORF1a:F3677-,S:H69-,S:V70-,S:Y144- |
| HP04068 | EPI_ISL_19 | B.1.1.7 | N:M1X,N:D | ORF1a:S3675-,ORF1a:G3676-,ORF1a:F3677-,ORF7a:S60-,ORF7a:T61-,ORF7a:Q62-,ORF7a:F63-,ORF7a |
| HP04071 | EPI_ISL_15 | B.1.526.1 | N:M1X,N:T | ORF1a:S3675-,ORF1a:G3676-,ORF1a:F3677- |
| HP04072 | EPI_ISL_15 | B.1.526.1 | N:M1X,N:T | ORF1a:S3675-,ORF1a:G3676-,ORF1a:F3677- |

| | | | | |
|---|---|---|---|---|
| HP04075 | EPI_ISL_15 | B.1.243 | N:S194L,N:I337V,ORF1a:V438A,ORF1a:G2118D,ORF1a:N2523T,ORF1b:P314L,ORF1b:A576V,ORF3a:T151I,ORF7a | |
| HP04076 | EPI_ISL_15 | B.1.1.7 | N:M1X,N:D | ORF1a:S3675-,ORF1a:G3676-,ORF1a:F3677-,S:H69-,S:V70-,S:Y144- |
| HP04077 | EPI_ISL_15 | B.1.1.7 | N:M1X,N:D | ORF1a:S3675-,ORF1a:G3676-,ORF1a:F3677-,S:H69-,S:V70-,S:Y144- |
| HP04080 | EPI_ISL_15 | B.1.1.519 | N:R203K,N:G204R,N:V350F,ORF1a:T224I,ORF1a:P959S,ORF1a:L1243P,ORF1a:T3255I,ORF1a:I3618V,ORF1a:T41 | |
| HP04124 | EPI_ISL_15 | B.1.1.7 | N:M1X,N:D | ORF1a:S3675-,ORF1a:G3676-,ORF1a:F3677-,S:H69-,S:V70-,S:Y144- |
| HP04125 | EPI_ISL_15 | B.1.1.7 | N:M1X,N:D | ORF1a:S3675-,ORF1a:G3676-,ORF1a:F3677-,S:H69-,S:V70-,S:Y144- |
| HP04126 | EPI_ISL_15 | B.1.526.1 | N:M1X,N:T | ORF1a:S3675-,ORF1a:G3676-,ORF1a:F3677-,S:Y144- |
| HP04127 | EPI_ISL_19 | B.1.1.7 | N:M1X,N:D | ORF1a:S3675-,ORF1a:G3676-,ORF1a:F3677-,ORF7a:S60-,ORF7a:T61-,ORF7a:Q62-,ORF7a:F63-,ORF7a |
| HP04128 | EPI_ISL_15 | B.1.1.7 | N:M1X,N:D | ORF1a:S3675-,ORF1a:G3676-,ORF1a:F3677-,S:H69-,S:V70-,S:Y144- |
| HP04149 | EPI_ISL_15 | B.1.1.7 | N:M1X,N:D | ORF1a:S3675-,ORF1a:G3676-,ORF1a:F3677-,S:H69-,S:V70-,S:Y144- |
| HP04150 | EPI_ISL_15 | B.1.1.7 | N:M1X,N:D | ORF1a:S3675-,ORF1a:G3676-,ORF1a:F3677-,S:H69-,S:V70-,S:Y144- |
| HP04154 | EPI_ISL_15 | B.1.526 | N:M1X,N:P | ORF1a:S3675-,ORF1a:G3676-,ORF1a:F3677- |
| HP04184 | EPI_ISL_15 | B.1.1.7 | N:M1X,N:D | ORF1a:S3675-,ORF1a:G3676-,ORF1a:F3677-,S:H69-,S:V70-,S:Y144- |
| HP04186 | EPI_ISL_15 | R.1 | M:F28L,N:M1X,N:S187L,N:R203K,N:G204R,N:Q418H,ORF1a:A2584T,ORF1b:P314L,ORF1b:V1144L,ORF1b:G1362 | |
| HP04187 | EPI_ISL_15 | B.1.1.7 | N:M1X,N:D | ORF1a:S3675-,ORF1a:G3676-,ORF1a:F3677-,S:H69-,S:V70-,S:Y144- |
| HP04188 | EPI_ISL_15 | B.1.561 | N:S194L,N:M234I,ORF1a:V751L,ORF1a:A1082V,ORF1a:I1274F,ORF1a:I1912V,ORF1a:K3353R,ORF1b:P314L,ORF1 | |
| HP04190 | EPI_ISL_16 | B.1.1.7 | N:M1X,N:D | ORF1a:S3675-,ORF1a:G3676-,ORF1a:F3677-,S:H69-,S:V70-,S:Y144- |
| HP04193 | EPI_ISL_19 | B.1.1.7 | N:M1X,N:D | ORF1a:S3675-,ORF1a:G3676-,ORF1a:F3677-,ORF7a:S60-,ORF7a:T61-,ORF7a:Q62-,ORF7a:F63-,ORF7a |
| HP04195 | EPI_ISL_16 | B.1.1.7 | N:M1X,N:D | ORF1a:S3675-,ORF1a:G3676-,ORF1a:F3677-,S:H69-,S:V70-,S:Y144- |
| HP04196 | EPI_ISL_16 | B.1.1.7 | N:M1X,N:D | ORF1a:S3675-,ORF1a:G3676-,ORF1a:F3677-,S:H69-,S:V70-,S:Y144- |
| HP04205 | EPI_ISL_16 | B.1.1.7 | N:M1X,N:D | ORF1a:S3675-,ORF1a:G3676-,ORF1a:F3677-,S:H69-,S:V70-,S:Y144- |
| HP04509 | EPI_ISL_16 | B.1.1.7 | N:M1X,N:D | ORF1a:S3675-,ORF1a:G3676-,ORF1a:F3677-,ORF3a:V256-,S:H69-,S:V70-,S:Y144- |
| HP04510 | EPI_ISL_16 | B.1.1.7 | N:M1X,N:D | ORF1a:S3675-,ORF1a:G3676-,ORF1a:F3677-,S:H69-,S:V70-,S:Y144- |
| HP04511 | EPI_ISL_16 | B.1.1.7 | N:M1X,N:D | ORF1a:S3675-,ORF1a:G3676-,ORF1a:F3677-,S:H69-,S:V70-,S:Y144- |
| HP04512 | EPI_ISL_16 | B.1.1.7 | N:M1X,N:D | ORF1a:S3675-,ORF1a:G3676-,ORF1a:F3677-,S:H69-,S:V70-,S:Y144- |
| HP04513 | EPI_ISL_16 | P.1 | N:P80R,N:F | ORF1a:S3675-,ORF1a:G3676-,ORF1a:F3677- |
| HP04514 | EPI_ISL_16 | B.1.1.7 | N:M1X,N:D | ORF1a:S3675-,ORF1a:G3676-,ORF1a:F3677-,S:H69-,S:V70-,S:Y144- |
| HP04515 | EPI_ISL_16 | B.1.1.7 | N:M1X,N:D | ORF1a:S3675-,ORF1a:G3676-,ORF1a:F3677-,S:H69-,S:V70-,S:Y144- |
| HP04516 | EPI_ISL_16 | B.1.1.7 | N:M1X,N:D | ORF1a:S3675-,ORF1a:G3676-,ORF1a:F3677-,S:H69-,S:V70-,S:Y144- |
| HP04517 | EPI_ISL_16 | B.1.1.7 | N:M1X,N:D | ORF1a:S3675-,ORF1a:G3676-,ORF1a:F3677-,S:H69-,S:V70-,S:Y144- |
| HP04520 | EPI_ISL_16 | B.1.1.7 | M:V23I,N:N | ORF1a:S3675-,ORF1a:G3676-,ORF1a:F3677-,S:H69-,S:V70-,S:Y144- |
| HP04521 | EPI_ISL_16 | B.1.351 | E:P71L,N:T. | ORF1a:S3675-,ORF1a:G3676-,ORF1a:F3677-,S:A243-,S:L244-,S:H245- |
| HP04522 | EPI_ISL_16 | B.1.243 | N:S194L,N:I337V,ORF1a:V438A,ORF1a:G2118Y,ORF1a:N2523T,ORF1b:P314L,ORF1b:A576V,ORF3a:T151I,S:D61 | |
| HP04523 | EPI_ISL_16 | B.1.1.7 | N:M1X,N:D | ORF1a:S3675-,ORF1a:G3676-,ORF1a:F3677-,S:H69-,S:V70-,S:Y144- |

| | | | | |
|---|---|---|---|---|
| HP04524 | EPI_ISL_16 | B.1.1.7 | N:M1X,N:D | ORF1a:S3675-,ORF1a:G3676-,ORF1a:F3677-,S:H69-,S:V70-,S:Y144- |
| HP04527 | EPI_ISL_16 | B.1.1.7 | N:M1X,N:D | ORF1a:S3675-,ORF1a:G3676-,ORF1a:F3677-,S:H69-,S:V70-,S:Y144- |
| HP04538 | EPI_ISL_16 | B.1.1.7 | N:M1X,N:D | ORF1a:S3675-,ORF1a:G3676-,ORF1a:F3677-,S:H69-,S:V70-,S:Y144- |
| HP04555 | EPI_ISL_16 | B.1.1.7 | N:M1X,N:D | ORF1a:S3675-,ORF1a:G3676-,ORF1a:F3677-,S:H69-,S:V70-,S:Y144- |
| HP04556 | EPI_ISL_16 | B.1.1.7 | N:M1X,N:D | ORF1a:S3675-,ORF1a:G3676-,ORF1a:F3677-,S:H69-,S:V70-,S:Y144- |
| HP04557 | EPI_ISL_16 | B.1.111 | N:T205I,N:K387R,ORF1a:I114T,ORF1a:M1312I,ORF1b:P314L,ORF1b:V1840F,ORF3a:G44E,ORF3a:Q57H,ORF3a:T | |
| HP04558 | EPI_ISL_16 | B.1.1.7 | N:M1X,N:D | ORF1a:S3675-,ORF1a:G3676-,ORF1a:F3677-,S:H69-,S:V70-,S:Y144- |
| HP04559 | EPI_ISL_16 | R.1 | M:F28L,N:M1X,N:S187L,N:R203K,N:G204R,N:Q418H,ORF1a:S1510F,ORF1a:T1854I,ORF1a:A2584T,ORF1b:P314L | |
| HP04560 | EPI_ISL_16 | B.1.1.7 | N:M1X,N:D | ORF1a:S3675-,ORF1a:G3676-,ORF1a:F3677-,S:H69-,S:V70-,S:Y144- |
| HP04561 | EPI_ISL_19 | B.1.1.7 | N:M1X,N:D | ORF1a:S3675-,ORF1a:G3676-,ORF1a:F3677-,S:H69-,S:V70-,S:Y144- |
| HP04562 | EPI_ISL_16 | B.1.1.7 | N:M1X,N:D | ORF1a:S3675-,ORF1a:G3676-,ORF1a:F3677-,S:H69-,S:V70-,S:Y144- |
| HP04563 | EPI_ISL_16 | B.1.1.7 | N:M1X,N:D | ORF1a:S3675-,ORF1a:G3676-,ORF1a:F3677-,S:H69-,S:V70-,S:Y144- |
| HP04569 | EPI_ISL_23 | B.1.1.7 | N:M1X,N:D | ORF1a:S3675-,ORF1a:G3676-,ORF1a:F3677-,S:H69-,S:V70-,S:Y144- |
| HP04570 | EPI_ISL_23 | B.1.1.7 | N:M1X,N:D | ORF1a:S3675-,ORF1a:G3676-,ORF1a:F3677-,S:H69-,S:V70-,S:Y144- |
| HP04578 | EPI_ISL_19 | B.1.1.318 | M:I82T,N:R | N:R209-,ORF1a:S3675-,ORF1a:G3676-,ORF1a:F3677-,ORF8:M1-,ORF8:K2-,S:Y144- |
| HP04579 | EPI_ISL_23 | B.1.1.7 | N:M1X,N:D | ORF1a:S3675-,ORF1a:G3676-,ORF1a:F3677-,S:H69-,S:V70-,S:Y144- |
| HP04581 | EPI_ISL_23 | B.1.1.519 | N:R203K,N | ORF3a:T14-,ORF3a:L15- |
| HP04582 | EPI_ISL_23 | B.1.427 | N:D144H,N:T205I,ORF1a:T265I,ORF1a:S3158T,ORF1b:P314L,ORF1b:P976L,ORF1b:D1183Y,ORF3a:Q57H,ORF3a: | |
| HP04583 | EPI_ISL_23 | B.1.526.1 | N:M1X,N:T | ORF1a:S3675-,ORF1a:G3676-,ORF1a:F3677- |
| HP04588 | EPI_ISL_23 | B.1.526.1 | N:M1X,N:T | ORF1a:S3675-,ORF1a:G3676-,ORF1a:F3677-,S:Y144- |
| HP04594 | EPI_ISL_23 | B.1.526 | N:M1X,N:R | ORF1a:S3675-,ORF1a:G3676-,ORF1a:F3677- |
| HP04596 | EPI_ISL_23 | B.1.1.7 | N:M1X,N:D | ORF1a:S3675-,ORF1a:G3676-,ORF1a:F3677-,S:H69-,S:V70-,S:Y144- |
| HP04597 | EPI_ISL_23 | B.1.526.1 | N:M1X,N:T | ORF1a:S3675-,ORF1a:G3676-,ORF1a:F3677- |
| HP04600 | EPI_ISL_23 | B.1.1.7 | N:M1X,N:D | ORF1a:S3675-,ORF1a:G3676-,ORF1a:F3677-,S:H69-,S:V70-,S:Y144- |
| HP04603 | pending | B.1.1.7 | N:M1X,N:D | ORF1a:S3675-,ORF1a:G3676-,ORF1a:F3677-,S:H69-,S:V70-,S:Y144- |
| HP04604 | EPI_ISL_23 | B.1.526.1 | N:M1X,N:T | ORF1a:S3675-,ORF1a:G3676-,ORF1a:F3677- |
| HP04659 | EPI_ISL_16 | B.1.1.1 | N:P13L,N:R | ORF1a:S3675-,ORF1a:G3676-,ORF1a:F3677-,ORF3a:L108- |
| HP04661 | EPI_ISL_16 | B.1.1.7 | N:M1X,N:D | ORF1a:S3675-,ORF1a:G3676-,ORF1a:F3677-,S:H69-,S:V70-,S:Y144- |
| HP04663 | EPI_ISL_16 | B.1.526 | N:M1X,N:D | ORF1a:S3675-,ORF1a:G3676-,ORF1a:F3677- |
| HP04664 | EPI_ISL_16 | B.1.526 | N:M1X,N:P | ORF1a:S3675-,ORF1a:G3676-,ORF1a:F3677- |
| HP04684 | EPI_ISL_19 | B.1.526 | N:M1X,N:P | ORF1a:S3675-,ORF1a:G3676-,ORF1a:F3677- |
| HP04721 | EPI_ISL_18 | B.1.351 | E:P71L,N:T | ORF1a:S3675-,ORF1a:G3676-,ORF1a:F3677-,S:A243-,S:L244-,S:H245- |
| HP04722 | EPI_ISL_18 | B.1.351 | E:P71L,N:T | ORF1a:S3675-,ORF1a:G3676-,ORF1a:F3677-,S:A243-,S:L244-,S:H245- |
| HP04723 | pending | B.1.526.1 | N:M1X,N:T | ORF1a:S3675-,ORF1a:G3676-,ORF1a:F3677-,ORF6:D61- |

| | | | | |
|---|---|---|---|---|
| HP04724 | EPI_ISL_18 | B.1.1.7 | N:M1X,N:D | ORF1a:S3675-,ORF1a:G3676-,ORF1a:F3677-,S:H69-,S:V70-,S:Y144- |
| HP04728 | EPI_ISL_18 | B.1.1.7 | N:M1X,N:D | ORF1a:S3675-,ORF1a:G3676-,ORF1a:F3677-,S:H69-,S:V70-,S:Y144- |
| HP04729 | EPI_ISL_18 | B.1.1.7 | N:M1X,N:D | ORF1a:S3675-,ORF1a:G3676-,ORF1a:F3677-,S:H69-,S:V70-,S:Y144- |
| HP04730 | EPI_ISL_18 | B.1.1.7 | N:M1X,N:D | ORF1a:S3675-,ORF1a:G3676-,ORF1a:F3677-,S:H69-,S:V70-,S:Y144- |
| HP04815 | EPI_ISL_21 | B.1 | N:M1X,N:S | ORF1a:S3675-,ORF1a:G3676-,ORF1a:F3677-,ORF3a:V256- |
| HP04816 | EPI_ISL_18 | B.1.1.7 | N:M1X,N:D | ORF1a:S3675-,ORF1a:G3676-,ORF1a:F3677-,S:H69-,S:V70-,S:Y144- |
| HP04819 | EPI_ISL_18 | B.1.1.7 | N:M1X,N:D | ORF1a:S3675-,ORF1a:G3676-,ORF1a:F3677-,S:H69-,S:V70-,S:Y144- |
| HP04820 | EPI_ISL_18 | B.1.1.7 | N:M1X,N:D | ORF1a:S3675-,ORF1a:G3676-,ORF1a:F3677-,S:H69-,S:V70-,S:Y144- |
| HP04821 | pending | B.1.526.1 | N:M1X,N:T | ORF1a:S3675-,ORF1a:G3676-,ORF1a:F3677-,ORF3a:P258-,ORF3a:V259- |
| HP04849 | EPI_ISL_18 | B.1.1.7 | N:M1X,N:D | ORF1a:S3675-,ORF1a:G3676-,ORF1a:F3677-,S:H69-,S:V70-,S:Y144- |
| HP04859 | EPI_ISL_18 | B.1.1.7 | N:M1X,N:D | ORF1a:S3675-,ORF1a:G3676-,ORF1a:F3677-,S:H69-,S:V70-,S:Y144- |
| HP04861 | EPI_ISL_18 | B.1.526.1 | N:M1X,N:T | ORF1a:S3675-,ORF1a:G3676-,ORF1a:F3677- |
| HP04864 | EPI_ISL_18 | B.1.1.7 | N:M1X,N:D | ORF1a:S3675-,ORF1a:G3676-,ORF1a:F3677-,S:H69-,S:V70-,S:Y144- |
| HP04865 | EPI_ISL_18 | B.1.1.7 | N:M1X,N:D | ORF1a:S3675-,ORF1a:G3676-,ORF1a:F3677-,S:H69-,S:V70-,S:Y144- |
| HP04866 | EPI_ISL_18 | B.1.1.7 | N:M1X,N:D | ORF1a:S3675-,ORF1a:G3676-,ORF1a:F3677-,S:H69-,S:V70-,S:Y144- |
| HP04867 | EPI_ISL_18 | B.1.1.7 | N:M1X,N:D | ORF1a:S3675-,ORF1a:G3676-,ORF1a:F3677-,S:H69-,S:V70-,S:Y144- |
| HP04868 | EPI_ISL_18 | B.1.1.7 | N:M1X,N:D | ORF1a:S3675-,ORF1a:G3676-,ORF1a:F3677-,S:H69-,S:V70-,S:Y144- |
| HP04870 | EPI_ISL_18 | B.1.1.7 | N:M1X,N:D | ORF1a:S3675-,ORF1a:G3676-,ORF1a:F3677-,S:H69-,S:V70-,S:Y144- |
| HP04874 | EPI_ISL_18 | B.1.1.7 | N:M1X,N:D | ORF1a:S3675-,ORF1a:G3676-,ORF1a:F3677-,S:H69-,S:V70-,S:Y144- |
| HP04875 | EPI_ISL_18 | B.1.1.7 | N:M1X,N:D | ORF1a:S3675-,ORF1a:G3676-,ORF1a:F3677-,S:H69-,S:V70-,S:Y144- |
| HP04876 | EPI_ISL_18 | B.1.526.1 | N:M1X,N:T | ORF1a:S3675-,ORF1a:G3676-,ORF1a:F3677- |
| HP04879 | EPI_ISL_18 | B.1.1.7 | N:M1X,N:D | ORF1a:S3675-,ORF1a:G3676-,ORF1a:F3677-,S:H69-,S:V70-,S:Y144- |
| HP04885 | EPI_ISL_18 | B.1.1.7 | N:M1X,N:D | ORF1a:S3675-,ORF1a:G3676-,ORF1a:F3677-,S:H69-,S:V70-,S:Y144- |
| HP04886 | EPI_ISL_18 | B.1.1.7 | N:M1X,N:D | ORF1a:S3675-,ORF1a:G3676-,ORF1a:F3677-,S:H69-,S:V70-,S:Y144- |
| HP04887 | EPI_ISL_18 | B.1.526.1 | N:M1X,N:T | ORF1a:S3675-,ORF1a:G3676-,ORF1a:F3677- |
| HP04888 | EPI_ISL_18 | B.1.1.7 | N:M1X,N:D | ORF1a:S3675-,ORF1a:G3676-,ORF1a:F3677-,S:H69-,S:V70-,S:Y144- |
| HP04915 | EPI_ISL_18 | B.1.526.1 | N:M1X,N:T | ORF1a:S3675-,ORF1a:G3676-,ORF1a:F3677- |
| HP04916 | EPI_ISL_18 | B.1.1.7 | N:M1X,N:D | ORF1a:S3675-,ORF1a:G3676-,ORF1a:F3677-,S:H69-,S:V70-,S:Y144- |
| HP04924 | pending | B.1.1.7 | M:G89S,N: | ORF1a:S3675-,ORF1a:G3676-,ORF1a:F3677-,S:H69-,S:V70-,S:Y144- |
| HP04925 | EPI_ISL_18 | B.1.1.7 | N:M1X,N:D | ORF1a:S3675-,ORF1a:G3676-,ORF1a:F3677-,S:H69-,S:V70-,S:Y144- |
| HP04926 | EPI_ISL_18 | B.1.1.7 | N:M1X,N:D | ORF1a:S3675-,ORF1a:G3676-,ORF1a:F3677-,S:H69-,S:V70-,S:Y144- |
| HP04927 | EPI_ISL_18 | B.1.1.7 | N:M1X,N:D | ORF1a:S3675-,ORF1a:G3676-,ORF1a:F3677-,S:H69-,S:V70-,S:Y144- |
| HP04928 | EPI_ISL_18 | B.1.1.7 | N:M1X,N:D | ORF1a:S3675-,ORF1a:G3676-,ORF1a:F3677-,S:H69-,S:V70-,S:Y144- |
| HP04929 | EPI_ISL_19 | B.1.1.7 | N:M1X,N:D | ORF1a:S3675-,ORF1a:G3676-,ORF1a:F3677-,S:H69-,S:V70-,S:Y144- |

| | | | | |
|---|---|---|---|---|
| HP04940 | EPI_ISL_18 | B.1.1.7 | N:M1X,N:D | ORF1a:S3675-,ORF1a:G3676-,ORF1a:F3677-,S:H69-,S:V70-,S:Y144- |
| HP04941 | EPI_ISL_19 | B.1.1.7 | N:M1X,N:D | ORF1a:S3675-,ORF1a:G3676-,ORF1a:F3677-,S:H69-,S:V70-,S:Y144- |
| HP04942 | EPI_ISL_19 | B.1.1.7 | N:M1X,N:D | ORF1a:H83-,ORF1a:V84-,ORF1a:M85-,ORF1a:V86-,ORF1a:E87-,ORF1a:S3675-,ORF1a:G3676-,ORF1a: |
| HP04967 | EPI_ISL_21 | B.1.1.7 | N:M1X,N:D | ORF1a:S3675-,ORF1a:G3676-,ORF1a:F3677-,S:H69-,S:V70-,S:Y144- |
| HP04986 | EPI_ISL_19 | B.1.525 | E:L21X,M:I | N:D3-,ORF1a:S3675-,ORF1a:G3676-,ORF1a:F3677-,ORF6:F2-,S:H69-,S:V70-,S:Y144- |
| HP04987 | EPI_ISL_19 | B.1.1.7 | N:M1X,N:D | ORF1a:S3675-,ORF1a:G3676-,ORF1a:F3677-,S:H69-,S:V70-,S:Y144- |
| HP04988 | EPI_ISL_19 | B.1.1.7 | N:M1X,N:D | ORF1a:S3675-,ORF1a:G3676-,ORF1a:F3677-,S:H69-,S:V70-,S:Y144- |
| HP04990 | EPI_ISL_19 | B.1.1.7 | N:M1X,N:D | ORF1a:S3675-,ORF1a:G3676-,ORF1a:F3677-,S:H69-,S:V70-,S:Y144- |
| HP04996 | EPI_ISL_19 | B.1.1.7 | N:M1X,N:D | ORF1a:S3675-,ORF1a:G3676-,ORF1a:F3677-,ORF7a:S60-,ORF7a:T61-,ORF7a:Q62-,ORF7a:F63-,ORF7a |
| HP04997 | EPI_ISL_19 | B.1.1.7 | N:M1X,N:D | ORF1a:S3675-,ORF1a:G3676-,ORF1a:F3677-,ORF7a:S60-,ORF7a:T61-,ORF7a:Q62-,ORF7a:F63-,ORF7a |
| HP04998 | EPI_ISL_19 | B.1.1.7 | N:M1X,N:D | ORF1a:S3675-,ORF1a:G3676-,ORF1a:F3677-,S:H69-,S:V70-,S:Y144- |
| HP05001 | EPI_ISL_19 | B.1.1.7 | N:M1X,N:D | ORF1a:S3675-,ORF1a:G3676-,ORF1a:F3677-,S:H69-,S:V70-,S:Y144- |
| HP05003 | EPI_ISL_19 | B.1.1.7 | N:M1X,N:D | ORF1a:S3675-,ORF1a:G3676-,ORF1a:F3677-,S:H69-,S:V70-,S:Y144- |
| HP05005 | EPI_ISL_19 | B.1.1.7 | N:M1X,N:D | ORF1a:S3675-,ORF1a:G3676-,ORF1a:F3677-,S:H69-,S:V70-,S:Y144- |
| HP05006 | EPI_ISL_19 | B.1.1.7 | N:M1X,N:D | ORF1a:S3675-,ORF1a:G3676-,ORF1a:F3677-,S:H69-,S:V70-,S:Y144- |
| HP05017 | EPI_ISL_19 | B.1.526.1 | N:M1X,N:T | ORF1a:S3675-,ORF1a:G3676-,ORF1a:F3677- |
| HP05018 | EPI_ISL_19 | B.1.1.7 | N:M1X,N:D | ORF1a:S3675-,ORF1a:G3676-,ORF1a:F3677-,S:H69-,S:V70-,S:Y144- |
| HP05020 | EPI_ISL_19 | B.1.1.7 | N:M1X,N:D | ORF1a:S3675-,ORF1a:G3676-,ORF1a:F3677-,S:H69-,S:V70-,S:Y144- |
| HP05021 | EPI_ISL_19 | B.1.1.7 | N:M1X,N:D | ORF1a:S3675-,ORF1a:G3676-,ORF1a:F3677-,S:H69-,S:V70-,S:Y144- |
| HP05026 | EPI_ISL_19 | B.1.620 | N:A220V,O | ORF1a:S3675-,ORF1a:G3676-,ORF1a:F3677-,ORF7b:L14-,S:H69-,S:V70-,S:Y144-,S:L242-,S:A243- |
| HP05027 | EPI_ISL_19 | B.1.1.7 | N:M1X,N:D | ORF1a:S3675-,ORF1a:G3676-,ORF1a:F3677-,S:H69-,S:V70-,S:Y144- |
| HP05034 | EPI_ISL_19 | B.1.1.7 | N:M1X,N:D | ORF1a:S3675-,ORF1a:G3676-,ORF1a:F3677-,S:H69-,S:V70-,S:Y144- |
| HP05043 | EPI_ISL_19 | B.1.1.7 | N:M1X,N:D | ORF1a:S3675-,ORF1a:G3676-,ORF1a:F3677-,S:H69-,S:V70-,S:Y144- |
| HP05046 | EPI_ISL_19 | B.1.1.7 | N:M1X,N:D | ORF1a:S3675-,ORF1a:G3676-,ORF1a:F3677-,S:H69-,S:V70-,S:Y144- |
| HP05050 | EPI_ISL_19 | B.1.1.7 | N:M1X,N:D | ORF1a:S3675-,ORF1a:G3676-,ORF1a:F3677-,S:H69-,S:V70-,S:Y144- |
| HP05051 | EPI_ISL_19 | B.1.1.7 | N:M1X,N:D | ORF1a:S3675-,ORF1a:G3676-,ORF1a:F3677-,S:H69-,S:V70-,S:Y144- |
| HP05052 | EPI_ISL_19 | B.1.1.7 | N:M1X,N:D | ORF1a:S3675-,ORF1a:G3676-,ORF1a:F3677-,S:H69-,S:V70-,S:Y144- |
| HP05053 | EPI_ISL_19 | B.1.526 | N:M1X,N:P | ORF1a:S3675-,ORF1a:G3676-,ORF1a:F3677- |
| HP05054 | EPI_ISL_19 | B.1.1.7 | M:H125Y,N | ORF1a:S3675-,ORF1a:G3676-,ORF1a:F3677-,S:H69-,S:V70-,S:Y144- |
| HP05056 | EPI_ISL_19 | B.1.427 | N:T205I,ORF1a:T265I,ORF1a:T2877I,ORF1a:S3158T,ORF1b:P314L,ORF1b:P976L,ORF1b:D1183Y,ORF1b:E1623D,( | |
| HP05065 | EPI_ISL_19 | B.1.1.7 | N:M1X,N:D | ORF1a:S3675-,ORF1a:G3676-,ORF1a:F3677-,S:H69-,S:V70-,S:Y144- |
| HP05071 | EPI_ISL_19 | P.1 | N:P80R,N:I | ORF1a:S3675-,ORF1a:G3676-,ORF1a:F3677- |
| HP05075 | EPI_ISL_19 | B.1.1.7 | N:M1X,N:D | ORF1a:S3675-,ORF1a:G3676-,ORF1a:F3677-,S:H69-,S:V70-,S:Y144- |
| HP05080 | pending | B.1.1.7 | N:M1X,N:D | ORF1a:S3675-,ORF1a:G3676-,ORF1a:F3677-,ORF8:S67-,S:H69-,S:V70-,S:Y144- |

| | | | | |
|---|---|---|---|---|
| HP05081 | EPI_ISL_19 | B.1.1.434 | N:R203K,N:G204R,ORF1a:T224I,ORF1a:I693V,ORF1a:Q998H,ORF1a:M1312I,ORF1a:L3776F,ORF1b:P314L,ORF1b | |
| HP05082 | EPI_ISL_19 | B.1.526.1 | N:M1X,N:T | ORF1a:S3675-,ORF1a:G3676-,ORF1a:F3677- |
| HP05085 | EPI_ISL_19 | B.1.1.7 | N:M1X,N:D | ORF1a:S3675-,ORF1a:G3676-,ORF1a:F3677-,S:H69-,S:V70-,S:Y144- |
| HP05087 | EPI_ISL_19 | B.1.1.7 | N:M1X,N:D | ORF1a:S3675-,ORF1a:G3676-,ORF1a:F3677-,S:H69-,S:V70-,S:Y144- |
| HP05088 | EPI_ISL_19 | B.1.1.7 | N:M1X,N:D | ORF1a:S3675-,ORF1a:G3676-,ORF1a:F3677-,S:H69-,S:V70-,S:Y144- |
| HP05089 | EPI_ISL_19 | B.1.526.1 | N:M1X,N:T | ORF1a:S3675-,ORF1a:G3676-,ORF1a:F3677- |
| HP05091 | EPI_ISL_19 | B.1.526.1 | N:M1X,N:T | ORF1a:S3675-,ORF1a:G3676-,ORF1a:F3677-,S:Y144- |
| HP05092 | EPI_ISL_19 | B.1.1.7 | N:M1X,N:D | ORF1a:S3675-,ORF1a:G3676-,ORF1a:F3677-,S:H69-,S:V70-,S:Y144- |
| HP05095 | EPI_ISL_19 | B.1.1.7 | N:M1X,N:D | ORF1a:S3675-,ORF1a:G3676-,ORF1a:F3677-,S:H69-,S:V70-,S:Y144- |
| HP05097 | EPI_ISL_19 | B.1.526 | N:M1X,N:D | ORF1a:S3675-,ORF1a:G3676-,ORF1a:F3677- |
| HP05099 | EPI_ISL_19 | B.1.1.7 | N:M1X,N:D | ORF1a:S3675-,ORF1a:G3676-,ORF1a:F3677-,S:H69-,S:V70-,S:Y144- |
| HP05104 | EPI_ISL_19 | B.1.1.7 | N:M1X,N:D | ORF1a:S3675-,ORF1a:G3676-,ORF1a:F3677-,S:H69-,S:V70-,S:Y144- |
| HP05118 | EPI_ISL_19 | B.1.1.7 | N:M1X,N:D | ORF1a:S3675-,ORF1a:G3676-,ORF1a:F3677-,S:H69-,S:V70-,S:Y144- |
| HP05126 | EPI_ISL_19 | B.1.1.7 | N:M1X,N:D | ORF1a:S3675-,ORF1a:G3676-,ORF1a:F3677-,S:H69-,S:V70-,S:Y144- |
| HP05127 | EPI_ISL_19 | B.1.1.7 | N:M1X,N:D | ORF1a:S3675-,ORF1a:G3676-,ORF1a:F3677-,S:H69-,S:V70-,S:Y144- |
| HP05128 | EPI_ISL_19 | B.1.1.7 | N:M1X,N:D | ORF1a:S3675-,ORF1a:G3676-,ORF1a:F3677-,S:H69-,S:V70-,S:Y144- |
| HP05161 | EPI_ISL_19 | B.1.526.1 | N:M1X,N:T | ORF1a:S3675-,ORF1a:G3676-,ORF1a:F3677- |
| HP05166 | EPI_ISL_19 | B.1.1.7 | N:M1X,N:D | ORF1a:S3675-,ORF1a:G3676-,ORF1a:F3677-,ORF7a:V108-,ORF7a:F109-,ORF7a:T111-,ORF7a:L112-,C |
| HP05169 | EPI_ISL_19 | B.1.1.7 | N:M1X,N:D | ORF1a:S3675-,ORF1a:G3676-,ORF1a:F3677-,S:H69-,S:V70-,S:Y144- |
| HP05174 | EPI_ISL_21 | B.1.1.7 | N:M1X,N:D | ORF1a:S3675-,ORF1a:G3676-,ORF1a:F3677-,ORF8:D119-,S:H69-,S:V70-,S:Y144- |
| HP05175 | EPI_ISL_19 | B.1.1.7 | N:M1X,N:D | ORF1a:S3675-,ORF1a:G3676-,ORF1a:F3677-,S:H69-,S:V70-,S:Y144- |
| HP05176 | pending | B.1.525 | E:L21X,M:I | N:D3-,ORF1a:S3675-,ORF1a:G3676-,ORF1a:F3677-,ORF6:F2-,S:H69-,S:V70-,S:Y144- |
| HP05177 | EPI_ISL_19 | B.1.234 | N:S194L,N:D371Y,N:T391I,N:S412N,ORF1a:R287S,ORF1a:I453T,ORF1a:V665I,ORF1a:T1682I,ORF1a:K2059R,ORF1 | |
| HP05178 | EPI_ISL_19 | B.1.526.1 | N:M1X,N:T | ORF1a:S3675-,ORF1a:G3676-,ORF1a:F3677- |
| HP05189 | EPI_ISL_19 | B.1.1.7 | N:M1X,N:D | ORF1a:S3675-,ORF1a:G3676-,ORF1a:F3677-,S:H69-,S:V70-,S:Y144- |
| HP05190 | EPI_ISL_19 | B.1.1.7 | N:M1X,N:D | ORF1a:S3675-,ORF1a:G3676-,ORF1a:F3677-,S:H69-,S:V70-,S:Y144- |
| HP05193 | EPI_ISL_19 | B.1.1.7 | N:M1X,N:D | ORF1a:S3675-,ORF1a:G3676-,ORF1a:F3677-,S:H69-,S:V70-,S:Y144- |
| HP05195 | EPI_ISL_19 | R.1 | M:F28L,N:N | S:H69-,S:V70-,S:Y144- |
| HP05198 | EPI_ISL_19 | B.1.1.7 | N:M1X,N:D | ORF1a:S3675-,ORF1a:G3676-,ORF1a:F3677-,S:H69-,S:V70-,S:Y144- |
| HP05199 | EPI_ISL_21 | B.1.1.7 | N:M1X,N:D | ORF1a:S3675-,ORF1a:G3676-,ORF1a:F3677-,S:H69-,S:V70-,S:Y144- |
| HP05200 | EPI_ISL_19 | B.1.1.7 | N:M1X,N:D | ORF1a:H83-,ORF1a:V84-,ORF1a:M85-,ORF1a:V86-,ORF1a:E87-,ORF1a:S3675-,ORF1a:G3676-,ORF1a: |
| HP05203 | EPI_ISL_19 | B.1.1.7 | N:M1X,N:D | ORF1a:S3675-,ORF1a:G3676-,ORF1a:F3677-,S:H69-,S:V70-,S:Y144- |
| HP05204 | EPI_ISL_19 | B.1.526.1 | N:M1X,N:T | ORF1a:S3675-,ORF1a:G3676-,ORF1a:F3677- |
| HP05210 | EPI_ISL_19 | B.1.1.7 | N:M1X,N:D | ORF1a:S3675-,ORF1a:G3676-,ORF1a:F3677-,S:H69-,S:V70-,S:Y144- |

| | | | | |
|---|---|---|---|---|
| HP05212 | EPI_ISL_19 | B.1.526.2 | N:M1X,N:P | ORF1a:S3675-,ORF1a:G3676-,ORF1a:F3677- |
| HP05250 | EPI_ISL_21 | B.1.1.7 | N:M1X,N:D | ORF1a:S3675-,ORF1a:G3676-,ORF1a:F3677-,S:H69-,S:V70-,S:Y144- |
| HP05287 | EPI_ISL_21 | B.1.1.7 | N:M1X,N:D | ORF1a:S3675-,ORF1a:G3676-,ORF1a:F3677-,S:H69-,S:V70-,S:Y144- |
| HP05289 | EPI_ISL_21 | B.1.1.7 | N:M1X,N:D | ORF1a:S3675-,ORF1a:G3676-,ORF1a:F3677-,S:H69-,S:V70-,S:Y144- |
| HP05299 | EPI_ISL_22 | B.1.1.7 | N:M1X,N:D | ORF1a:S3675-,ORF1a:G3676-,ORF1a:F3677-,S:H69-,S:V70-,S:Y144- |
| HP05300 | EPI_ISL_21 | B.1.1.7 | ORF1a:T10 | ORF1a:S3675-,ORF1a:G3676-,ORF1a:F3677-,S:H69-,S:V70-,S:Y144- |
| HP05302 | EPI_ISL_22 | B.1.1.7 | N:M1X,N:D | ORF1a:S3675-,ORF1a:G3676-,ORF1a:F3677-,S:H69-,S:V70-,S:Y144- |
| HP05303 | EPI_ISL_22 | B.1.1.7 | N:M1X,N:D | ORF1a:S3675-,ORF1a:G3676-,ORF1a:F3677-,S:H69-,S:V70-,S:Y144- |
| HP05307 | EPI_ISL_21 | B.1.1.7 | N:M1X,N:D | ORF1a:S3675-,ORF1a:G3676-,ORF1a:F3677-,S:H69-,S:V70-,S:Y144- |
| HP05308 | EPI_ISL_21 | B.1.1.7 | N:M1X,N:D | ORF1a:S3675-,ORF1a:G3676-,ORF1a:F3677-,S:H69-,S:V70-,S:Y144- |
| HP05309 | EPI_ISL_21 | B.1.1.7 | N:M1X,N:D | ORF1a:S3675-,ORF1a:G3676-,ORF1a:F3677-,S:H69-,S:V70-,S:Y144- |
| HP05310 | EPI_ISL_22 | B.1.1.7 | N:M1X,N:D | ORF1a:S3675-,ORF1a:G3676-,ORF1a:F3677-,S:H69-,S:V70-,S:Y144- |
| HP05311 | EPI_ISL_22 | B.1.1.7 | N:M1X,N:D | ORF1a:S3675-,ORF1a:G3676-,ORF1a:F3677-,S:H69-,S:V70-,S:Y144- |
| HP05312 | EPI_ISL_22 | B.1.1.7 | N:M1X,N:D | ORF1a:S3675-,ORF1a:G3676-,ORF1a:F3677-,S:H69-,S:V70-,S:Y144- |
| HP05319 | EPI_ISL_22 | B.1.1.7 | N:M1X,N:D | ORF1a:S3675-,ORF1a:G3676-,ORF1a:F3677-,S:H69-,S:V70-,S:Y144- |
| HP05325 | EPI_ISL_22 | B.1.1.7 | N:M1X,N:D | ORF1a:S3675-,ORF1a:G3676-,ORF1a:F3677-,S:H69-,S:V70-,S:Y144- |
| HP05326 | pending | B.1.1.7 | N:M1X,N:D | ORF1a:S3675-,ORF1a:G3676-,ORF1a:F3677-,ORF7a:S60-,ORF7a:T61-,ORF7a:Q62-,ORF7a:F63-,ORF7a |
| HP05327 | EPI_ISL_22 | B.1.1.7 | N:M1X,N:D | ORF1a:S3675-,ORF1a:G3676-,ORF1a:F3677-,S:H69-,S:V70-,S:Y144- |
| HP05332 | EPI_ISL_22 | B.1.1.7 | N:M1X,N:D | ORF1a:S3675-,ORF1a:G3676-,ORF1a:F3677-,S:H69-,S:V70-,S:Y144-,S:L242-,S:A243-,S:L244-,S:H245-,S |
| HP05334 | EPI_ISL_22 | B.1.1.7 | N:M1X,N:D | ORF1a:S3675-,ORF1a:G3676-,ORF1a:F3677-,S:H69-,S:V70-,S:Y144- |
| HP05348 | EPI_ISL_22 | B.1.1.7 | N:M1X,N:D | ORF1a:S3675-,ORF1a:G3676-,ORF1a:F3677-,S:H69-,S:V70-,S:Y144- |
| HP05349 | EPI_ISL_22 | B.1.1.7 | N:M1X,N:D | ORF1a:S3675-,ORF1a:G3676-,ORF1a:F3677-,S:H69-,S:V70-,S:Y144- |
| HP05356 | EPI_ISL_22 | B.1.1.7 | N:M1X,N:D | ORF1a:S3675-,ORF1a:G3676-,ORF1a:F3677-,S:H69-,S:V70-,S:Y144- |
| HP05357 | EPI_ISL_22 | B.1.526 | N:M1X,N:P | ORF1a:S3675-,ORF1a:G3676-,ORF1a:F3677- |
| HP05359 | EPI_ISL_22 | B.1 | N:M1X,N:T | ORF1a:S3675-,ORF1a:G3676-,ORF1a:F3677- |
| HP05361 | EPI_ISL_22 | B.1.1.7 | N:M1X,N:D | ORF1a:S3675-,ORF1a:G3676-,ORF1a:F3677-,ORF3a:V256-,S:H69-,S:V70-,S:Y144- |
| HP05363 | EPI_ISL_22 | B.1.1.7 | N:M1X,N:D | ORF1a:S3675-,ORF1a:G3676-,ORF1a:F3677-,S:H69-,S:V70-,S:Y144-,S:L242-,S:A243-,S:L244-,S:H245-,S |
| HP05395 | EPI_ISL_23 | B.1.1.7 | N:M1X,N:D | ORF1a:S3675-,ORF1a:G3676-,ORF1a:F3677-,S:H69-,S:V70-,S:Y144- |
| HP05396 | pending | B.1.1.7 | N:M1X,N:D | ORF1a:S3675-,ORF1a:G3676-,ORF1a:F3677-,S:H69-,S:V70-,S:Y144- |
| HP05398 | EPI_ISL_23 | B.1.1.7 | N:M1X,N:D | ORF1a:S3675-,ORF1a:G3676-,ORF1a:F3677-,S:H69-,S:V70-,S:Y144- |
| HP05399 | pending | B.1.1.7 | N:M1X,N:D | ORF1a:S3675-,ORF1a:G3676-,ORF1a:F3677-,S:H69-,S:V70-,S:Y144- |
| HP05403 | EPI_ISL_23 | B.1.1.7 | M:A69S,N:I | ORF1a:S3675-,ORF1a:G3676-,ORF1a:F3677-,S:H69-,S:V70-,S:Y144- |
| HP05407 | EPI_ISL_23 | B.1.1.7 | N:M1X,N:D | ORF1a:S3675-,ORF1a:G3676-,ORF1a:F3677-,S:H69-,S:V70-,S:Y144- |
| HP05408 | EPI_ISL_23 | B.1.1.7 | N:M1X,N:D | ORF1a:S3675-,ORF1a:G3676-,ORF1a:F3677-,S:H69-,S:V70-,S:Y144- |

| | | | | |
|---|---|---|---|---|
| HP05411 | EPI_ISL_23 | B.1.1.7 | N:M1X,N:D | ORF1a:S3675-,ORF1a:G3676-,ORF1a:F3677-,S:H69-,S:V70-,S:Y144- |
| HP05417 | EPI_ISL_23 | B.1.1.7 | N:M1X,N:D | ORF1a:S3675-,ORF1a:G3676-,ORF1a:F3677-,S:H69-,S:V70-,S:Y144- |
| HP05422 | EPI_ISL_23 | B.1.526.3 | N:M1X,N:P | ORF1a:S3675-,ORF1a:G3676-,ORF1a:F3677- |
| HP05424 | EPI_ISL_23 | B.1.1.7 | N:M1X,N:D | ORF1a:S3675-,ORF1a:G3676-,ORF1a:F3677-,S:H69-,S:V70-,S:Y144- |
| HP05425 | EPI_ISL_23 | B.1.1.7 | N:M1X,N:D | ORF1a:S3675-,ORF1a:G3676-,ORF1a:F3677-,S:H69-,S:V70-,S:Y144- |
| HP05429 | EPI_ISL_23 | B.1.1.7 | N:M1X,N:D | ORF1a:S3675-,ORF1a:G3676-,ORF1a:F3677-,S:H69-,S:V70-,S:Y144- |
| HP05432 | EPI_ISL_23 | B.1.1.7 | N:M1X,N:D | ORF1a:S3675-,ORF1a:G3676-,ORF1a:F3677-,S:H69-,S:V70-,S:Y144- |
| HP05433 | EPI_ISL_23 | B.1.1.7 | N:M1X,N:D | ORF1a:S3675-,ORF1a:G3676-,ORF1a:F3677-,S:H69-,S:V70-,S:Y144- |
| HP05434 | EPI_ISL_23 | B.1.1.7 | N:M1X,N:D | ORF1a:S3675-,ORF1a:G3676-,ORF1a:F3677-,S:H69-,S:V70-,S:Y144- |
| HP05441 | EPI_ISL_23 | B.1.1.7 | N:M1X,N:D | ORF1a:S3675-,ORF1a:G3676-,ORF1a:F3677-,S:H69-,S:V70-,S:Y144- |
| HP05443 | EPI_ISL_23 | B.1.1.7 | N:M1X,N:D | ORF1a:S3675-,ORF1a:G3676-,ORF1a:F3677-,S:H69-,S:V70-,S:Y144- |
| HP05448 | EPI_ISL_23 | B.1.1.7 | N:M1X,N:D | ORF1a:S3675-,ORF1a:G3676-,ORF1a:F3677-,S:H69-,S:V70-,S:Y144- |
| HP05449 | EPI_ISL_23 | B.1.1.7 | N:M1X,N:D | ORF1a:S3675-,ORF1a:G3676-,ORF1a:F3677-,S:H69-,S:V70-,S:Y144- |
| HP05452 | EPI_ISL_23 | B.1.526.1 | N:M1X,N:T | ORF1a:S3675-,ORF1a:G3676-,ORF1a:F3677-,S:Y144- |
| HP05455 | EPI_ISL_23 | B.1.1.7 | N:M1X,N:D | ORF1a:S3675-,ORF1a:G3676-,ORF1a:F3677-,S:H69-,S:V70-,S:Y144- |
| HP05456 | EPI_ISL_23 | B.1.1.7 | N:M1X,N:D | ORF1a:S3675-,ORF1a:G3676-,ORF1a:F3677-,S:H69-,S:V70-,S:Y144- |
| HP05458 | EPI_ISL_23 | B.1.427 | N:A90S,N:T205I,ORF1a:T265I,ORF1a:A1352V,ORF1a:S3158T,ORF1a:G3546S,ORF1b:P314L,ORF1b:P976L,ORF1b: | |
| HP05462 | EPI_ISL_23 | B.1.1.7 | N:M1X,N:D | ORF1a:S3675-,ORF1a:G3676-,ORF1a:F3677-,S:H69-,S:V70-,S:Y144- |
| HP05472 | EPI_ISL_23 | B.1.1.7 | N:M1X,N:D | ORF1a:S3675-,ORF1a:G3676-,ORF1a:F3677-,S:H69-,S:V70-,S:Y144- |
| HP05511 | EPI_ISL_21 | B.1.1.7 | N:M1X,N:D | ORF1a:S3675-,ORF1a:G3676-,ORF1a:F3677-,S:H69-,S:V70-,S:Y144- |
| HP05537 | EPI_ISL_22 | B.1.1.7 | N:M1X,N:D | ORF1a:S3675-,ORF1a:G3676-,ORF1a:F3677-,ORF8:L60-,S:H69-,S:V70-,S:Y144- |
| HP05539 | EPI_ISL_21 | B.1.526.1 | N:M1X,N:T | ORF1a:S3675-,ORF1a:G3676-,ORF1a:F3677-,S:Y144- |
| HP05540 | EPI_ISL_21 | B.1.1.7 | E:F26L,N:M | ORF1a:S3675-,ORF1a:G3676-,ORF1a:F3677-,S:H69-,S:V70-,S:Y144- |
| HP05543 | EPI_ISL_21 | B.1.1.7 | N:M1X,N:D | ORF1a:S3675-,ORF1a:G3676-,ORF1a:F3677-,S:H69-,S:V70-,S:Y144- |
| HP05550 | EPI_ISL_21 | B.1.526 | N:M1X,N:P | ORF1a:S3675-,ORF1a:G3676-,ORF1a:F3677- |
| HP05560 | EPI_ISL_21 | B.1.1.7 | N:M1X,N:D | ORF1a:S3675-,ORF1a:G3676-,ORF1a:F3677-,S:H69-,S:V70-,S:Y144- |
| HP05563 | EPI_ISL_22 | B.1.1.7 | N:M1X,N:D | ORF1a:S3675-,ORF1a:G3676-,ORF1a:F3677-,S:H69-,S:V70-,S:Y144- |
| HP05567 | EPI_ISL_21 | B.1.1.7 | N:M1X,N:D | ORF1a:S3675-,ORF1a:G3676-,ORF1a:F3677-,S:H69-,S:V70-,S:Y144- |
| HP05569 | EPI_ISL_21 | B.1.1.7 | N:M1X,N:D | ORF1a:S3675-,ORF1a:G3676-,ORF1a:F3677-,S:H69-,S:V70-,S:Y144- |
| HP05571 | EPI_ISL_21 | B.1.526.1 | N:M1X,N:T | ORF1a:S3675-,ORF1a:G3676-,ORF1a:F3677- |
| HP05572 | EPI_ISL_22 | B.1.1.7 | N:M1X,N:D | ORF1a:S3675-,ORF1a:G3676-,ORF1a:F3677-,S:H69-,S:V70-,S:Y144- |
| HP05573 | EPI_ISL_21 | B.1.1.7 | N:M1X,N:D | ORF1a:S3675-,ORF1a:G3676-,ORF1a:F3677-,S:H69-,S:V70-,S:Y144- |
| HP05574 | EPI_ISL_21 | B.1.1.7 | N:M1X,N:D | ORF1a:S3675-,ORF1a:G3676-,ORF1a:F3677-,S:H69-,S:V70-,S:Y144- |
| HP05575 | EPI_ISL_21 | B.1.1.7 | N:M1X,N:D | ORF1a:S3675-,ORF1a:G3676-,ORF1a:F3677-,S:H69-,S:V70-,S:Y144- |

| | | | | |
|---|---|---|---|---|
| HP05589 | EPI_ISL_21 | B.1.526.1 | N:M1X,N:T | ORF1a:S3675-,ORF1a:G3676-,ORF1a:F3677-,S:Y144- |
| HP05590 | EPI_ISL_21 | B.1.1.7 | N:M1X,N:D | ORF1a:S3675-,ORF1a:G3676-,ORF1a:F3677-,S:H69-,S:V70-,S:Y144- |
| HP05591 | EPI_ISL_21 | B.1.526.1 | N:M1X,N:T | ORF1a:S3675-,ORF1a:G3676-,ORF1a:F3677- |
| HP05593 | EPI_ISL_21 | B.1.526.1 | N:M1X,N:T | ORF1a:S3675-,ORF1a:G3676-,ORF1a:F3677-,S:Y144- |
| HP05594 | EPI_ISL_21 | B.1.1.7 | N:M1X,N:D | ORF1a:S3675-,ORF1a:G3676-,ORF1a:F3677-,S:H69-,S:V70-,S:Y144- |
| HP05597 | EPI_ISL_22 | B.1.1.7 | N:M1X,N:D | ORF1a:S3675-,ORF1a:G3676-,ORF1a:F3677-,S:H69-,S:V70-,S:Y144- |
| HP05598 | EPI_ISL_22 | B.1.1.7 | N:M1X,N:D | ORF1a:S3675-,ORF1a:G3676-,ORF1a:F3677-,S:H69-,S:V70-,S:Y144- |
| HP05599 | EPI_ISL_22 | B.1 | N:M1X,N:S | ORF1a:L3674-,ORF3a:V256- |
| HP05644 | pending | B.1.1.7 | N:M1X,N:D | ORF1a:S3675-,ORF1a:G3676-,ORF1a:F3677-,ORF3a:V256-,S:H69-,S:V70-,S:Y144- |
| HP05645 | pending | B.1.1.7 | N:M1X,N:D | ORF1a:S3675-,ORF1a:G3676-,ORF1a:F3677-,ORF3a:V256-,S:H69-,S:V70-,S:Y144- |
| HP05648 | EPI_ISL_23 | B.1.1.7 | N:M1X,N:D | ORF1a:S3675-,ORF1a:G3676-,ORF1a:F3677-,S:H69-,S:V70-,S:Y144- |
| HP05653 | EPI_ISL_23 | B.1.1.7 | N:M1X,N:D | ORF1a:S3675-,ORF1a:G3676-,ORF1a:F3677-,S:H69-,S:V70-,S:Y144- |
| HP05654 | EPI_ISL_23 | B.1.1.7 | N:M1X,N:D | ORF1a:S3675-,ORF1a:G3676-,ORF1a:F3677-,S:H69-,S:V70-,S:Y144- |
| HP05655 | EPI_ISL_23 | B.1.1.7 | N:M1X,N:D | ORF1a:S3675-,ORF1a:G3676-,ORF1a:F3677-,S:H69-,S:V70-,S:Y144- |
| HP05659 | EPI_ISL_23 | P.1 | N:P80R,N:I | ORF1a:S3675-,ORF1a:G3676-,ORF1a:F3677- |
| HP05660 | EPI_ISL_23 | B.1.617.2 | M:I82T,N:N | ORF8:D119-,ORF8:F120-,S:F157-,S:R158- |
| HP05661 | EPI_ISL_23 | B.1.1 | N:M1X,N:R | ORF8:D119-,ORF8:F120-,S:H69-,S:V70-,S:Y144-,S:F157-,S:R158- |
| HP05676 | EPI_ISL_23 | B.1.526.1 | N:M1X,N:T | ORF1a:S3675-,ORF1a:G3676-,ORF1a:F3677- |
| HP05678 | EPI_ISL_23 | B.1.1.7 | N:M1X,N:D | ORF1a:S3675-,ORF1a:G3676-,ORF1a:F3677-,S:H69-,S:V70-,S:Y144- |
| HP05679 | EPI_ISL_23 | B.1.1.7 | N:M1X,N:D | ORF1a:S3675-,ORF1a:G3676-,ORF1a:F3677-,S:H69-,S:V70-,S:Y144- |
| HP05680 | EPI_ISL_23 | B.1.1.7 | N:M1X,N:D | ORF1a:S3675-,ORF1a:G3676-,ORF1a:F3677-,S:H69-,S:V70-,S:Y144- |
| HP05681 | EPI_ISL_23 | B.1.1.7 | N:M1X,N:D | ORF1a:S3675-,ORF1a:G3676-,ORF1a:F3677-,S:H69-,S:V70-,S:Y144- |
| HP05682 | EPI_ISL_23 | B.1.1.7 | N:M1X,N:D | ORF1a:S3675-,ORF1a:G3676-,ORF1a:F3677-,S:H69-,S:V70-,S:Y144- |
| HP05684 | EPI_ISL_23 | B.1.1.7 | N:M1X,N:D | ORF1a:S3675-,ORF1a:G3676-,ORF1a:F3677-,S:H69-,S:V70-,S:Y144- |
| HP05693 | EPI_ISL_23 | B.1.1.7 | N:M1X,N:D | ORF1a:S3675-,ORF1a:G3676-,ORF1a:F3677-,S:H69-,S:V70-,S:Y144- |
| HP05694 | EPI_ISL_23 | B.1.1.7 | N:M1X,N:D | ORF1a:S3675-,ORF1a:G3676-,ORF1a:F3677-,S:H69-,S:V70-,S:Y144- |
| HP05701 | EPI_ISL_23 | B.1.1.7 | N:M1X,N:D | ORF1a:S3675-,ORF1a:G3676-,ORF1a:F3677-,S:H69-,S:V70-,S:Y144- |
| HP05705 | EPI_ISL_23 | B.1.526.1 | N:M1X,N:T | ORF1a:S3675-,ORF1a:G3676-,ORF1a:F3677- |
| HP05723 | EPI_ISL_23 | B.1.526 | N:M1X,N:P | ORF1a:S3675-,ORF1a:G3676-,ORF1a:F3677- |
| HP05724 | EPI_ISL_23 | B.1.1.7 | N:M1X,N:D | ORF1a:S3675-,ORF1a:G3676-,ORF1a:F3677-,S:H69-,S:V70-,S:Y144- |
| HP05767 | EPI_ISL_22 | B.1.1.7 | N:M1X,N:D | ORF1a:S3675-,ORF1a:G3676-,ORF1a:F3677-,S:H69-,S:V70-,S:Y144- |
| HP05768 | EPI_ISL_22 | B.1.1.7 | N:M1X,N:D | ORF1a:S3675-,ORF1a:G3676-,ORF1a:F3677-,S:H69-,S:V70-,S:Y144- |
| HP05772 | EPI_ISL_22 | B.1.1.7 | N:M1X,N:D | ORF1a:S3675-,ORF1a:G3676-,ORF1a:F3677-,S:H69-,S:V70-,S:Y144- |
| HP05774 | EPI_ISL_22 | B.1.1.7 | N:M1X,N:D | ORF1a:S3675-,ORF1a:G3676-,ORF1a:F3677-,S:H69-,S:V70-,S:Y144- |

| | | | |
|---|---|---|---|
| HP05775 | EPI_ISL_22 | A.23.1 | N:S2Y,N:S202N,N:R203K,ORF1a:T350N,ORF1a:T654I,ORF1a:T2846I,ORF1a:L3338F,ORF1a:M3655I,ORF1a:L3667 |
| HP05776 | EPI_ISL_22 | B.1.1.7 | N:M1X,N:D ORF1a:S3675-,ORF1a:G3676-,ORF1a:F3677-,S:H69-,S:V70-,S:Y144- |
| HP05777 | EPI_ISL_22 | B.1.1.7 | N:M1X,N:D ORF1a:S3675-,ORF1a:G3676-,ORF1a:F3677-,S:H69-,S:V70-,S:Y144- |
| HP05797 | EPI_ISL_22 | B.1.1.7 | N:M1X,N:D ORF1a:S3675-,ORF1a:G3676-,ORF1a:F3677-,S:H69-,S:V70-,S:Y144- |
| HP05802 | EPI_ISL_22 | B.1.1.7 | N:M1X,N:D ORF1a:S3675-,ORF1a:G3676-,ORF1a:F3677-,S:H69-,S:V70-,S:Y144- |
| HP05807 | EPI_ISL_22 | B.1.1.7 | N:M1X,N:D ORF1a:S3675-,ORF1a:G3676-,ORF1a:F3677-,S:H69-,S:V70-,S:Y144- |
| HP05810 | EPI_ISL_22 | B.1.1.7 | N:M1X,N:D ORF1a:S3675-,ORF1a:G3676-,ORF1a:F3677-,S:H69-,S:V70-,S:Y144- |
| HP05817 | EPI_ISL_22 | B.1.1.7 | N:R203K,N ORF1a:S3675-,ORF1a:G3676-,ORF1a:F3677-,S:H69-,S:V70-,S:Y144- |
| HP05818 | EPI_ISL_22 | B.1.1.7 | N:M1X,N:D ORF1a:S3675-,ORF1a:G3676-,ORF1a:F3677-,S:H69-,S:V70-,S:Y144- |
| HP05824 | EPI_ISL_22 | B.1.1.7 | N:M1X,N:D ORF1a:S3675-,ORF1a:G3676-,ORF1a:F3677-,S:H69-,S:V70-,S:Y144- |
| HP06072 | EPI_ISL_23 | B.1.1.7 | N:M1X,N:D ORF1a:S3675-,ORF1a:G3676-,ORF1a:F3677-,S:H69-,S:V70-,S:Y144- |
| HP06073 | EPI_ISL_23 | B.1.1.7 | N:M1X,N:D ORF1a:S3675-,ORF1a:G3676-,ORF1a:F3677-,S:H69-,S:V70-,S:Y144- |
| HP06074 | EPI_ISL_23 | B.1.1.7 | N:M1X,N:D ORF1a:S3675-,ORF1a:G3676-,ORF1a:F3677-,S:H69-,S:V70-,S:Y144- |
| HP06075 | EPI_ISL_23 | B.1.1.7 | N:M1X,N:D ORF1a:S3675-,ORF1a:G3676-,ORF1a:F3677-,S:H69-,S:V70-,S:Y144- |
| HP06078 | EPI_ISL_23 | B.1.1.7 | N:M1X,N:D ORF1a:S3675-,ORF1a:G3676-,ORF1a:F3677-,S:H69-,S:V70-,S:Y144- |
| HP06111 | EPI_ISL_23 | B.1.1.7 | N:M1X,N:D ORF1a:S3675-,ORF1a:G3676-,ORF1a:F3677-,S:H69-,S:V70-,S:Y144- |
| HP06113 | EPI_ISL_23 | B.1.1.7 | N:M1X,N:D ORF1a:S3675-,ORF1a:G3676-,ORF1a:F3677-,S:H69-,S:V70-,S:Y144- |
| HP06114 | EPI_ISL_23 | B.1.1.7 | N:M1X,N:D ORF1a:S3675-,ORF1a:G3676-,ORF1a:F3677-,S:H69-,S:V70-,S:Y144- |
| HP06235 | pending | B.1.2 | M:A83T,N:P67S,N:A414S,ORF1a:T760I,ORF1a:I2076T,ORF1a:M2606I,ORF1a:L3352F,ORF1a:V4073I,ORF1b:K82R |
| HP06243 | pending | B.1.1.7 | N:M1X,N:D ORF1a:S3675-,ORF1a:G3676-,ORF1a:F3677-,S:H69-,S:V70-,S:Y144- |
| HP06245 | pending | B.1.1.7 | N:M1X,N:D ORF1a:S3675-,ORF1a:G3676-,ORF1a:F3677-,ORF6:K23-,ORF6:V24-,ORF6:S25-,ORF6:I26-,ORF6:W27- |
| HP06247 | pending | B.1.1.7 | N:M1X,N:D ORF1a:S3675-,ORF1a:G3676-,ORF1a:F3677-,S:H69-,S:V70-,S:Y144- |
| HP06258 | pending | B.1.1.7 | N:M1X,N:D ORF1a:S3675-,ORF1a:G3676-,ORF1a:F3677-,S:H69-,S:V70-,S:Y144- |
| HP06261 | pending | B.1.1.7 | N:M1X,N:D ORF1a:S3675-,ORF1a:G3676-,ORF1a:F3677-,S:H69-,S:V70-,S:Y144- |
| HP06262 | pending | B.1.1.7 | N:M1X,N:D ORF1a:S3675-,ORF1a:G3676-,ORF1a:F3677-,S:H69-,S:V70-,S:Y144- |
| HP06271 | pending | B.1.1.7 | N:M1X,N:D ORF1a:S3675-,ORF1a:G3676-,ORF1a:F3677-,S:H69-,S:V70-,S:Y144- |
| HP06272 | pending | B.1.1.7 | N:M1X,N:D ORF1a:S3675-,ORF1a:G3676-,ORF1a:F3677-,S:H69-,S:V70-,S:Y144- |
| HP06273 | pending | B.1.526.2 | N:M1X,ORI ORF1a:S3675-,ORF1a:G3676-,ORF1a:F3677- |
| HP06274 | pending | B.1.525 | E:L21X,M:I N:D3-,ORF1a:S3675-,ORF1a:G3676-,ORF1a:F3677-,ORF6:F2-,S:H69-,S:V70-,S:Y144- |
| HP06276 | pending | B.1.1.7 | N:M1X,N:D ORF1a:S3675-,ORF1a:G3676-,ORF1a:F3677-,S:H69-,S:V70-,S:Y144- |
| HP06281 | pending | B.1.1.7 | N:M1X,N:D ORF1a:S3675-,ORF1a:G3676-,ORF1a:F3677-,S:H69-,S:V70-,S:Y144- |
| HP06302 | pending | B.1.2 | N:P67S,N:P199L,N:H300Y,ORF1a:D203N,ORF1a:T265I,ORF1a:M2606I,ORF1a:L3352F,ORF1a:R3993L,ORF1b:P31 |

Supplemental Table 4

| | outcome | Lineage | c | p value | dof | expected |
|---|---|---|---|---|---|---|
| **0** | Control | B.1.1.486 | 0.768827 | 0.38058 | 1 | [[6.48293963e+01 1.70603675e-01] [3.15170604e+02 8.29396325e-01]] |
| **1** | Control | B.1.1.7 | 2.337644 | 0.12628 | 1 | [[ 15.69553806 49.30446194] [ 76.30446194 239.69553806]] |
| **2** | Control | B.1.2 | 0.271578 | 0.602275 | 1 | [[ 63.97637795 1.02362205] [311.02362205 4.97637795]] |
| **3** | Control | B.1.214.2 | 0.768827 | 0.38058 | 1 | [[6.48293963e+01 1.70603675e-01] [3.15170604e+02 8.29396325e-01]] |
| **4** | Control | B.1.234 | 0.768827 | 0.38058 | 1 | [[6.48293963e+01 1.70603675e-01] [3.15170604e+02 8.29396325e-01]] |
| **5** | Control | B.1.243 | 0.000331 | 0.985479 | 1 | [[ 64.48818898 0.51181102] [313.51181102 2.48818898]] |
| **6** | Control | B.1.311 | 0.768827 | 0.38058 | 1 | [[6.48293963e+01 1.70603675e-01] [3.15170604e+02 8.29396325e-01]] |
| **7** | Control | B.1.351 | 0.096163 | 0.756484 | 1 | [[ 63.80577428 1.19422572] [310.19422572 5.80577428]] |
| **8** | Control | B.1.427 | 0.000331 | 0.985479 | 1 | [[ 64.48818898 0.51181102] [313.51181102 2.48818898]] |
| **9** | Control | B.1.525 | 0.000331 | 0.985479 | 1 | [[ 64.48818898 0.51181102] [313.51181102 2.48818898]] |
| **10** | Control | B.1.526 | 8.084124 | 0.004465 | 1 | [[ 61.92913386 3.07086614] [301.07086614 14.92913386]] |
| **11** | Control | B.1.526.1 | 1.83817 | 0.175166 | 1 | [[ 58.51706037 6.48293963] [284.48293963 31.51706037]] |
| **12** | Control | B.1.526.3 | 0.089571 | 0.764724 | 1 | [[ 64.65879265 0.34120735] [314.34120735 1.65879265]] |
| **13** | Control | B.1.596 | 0.768827 | 0.38058 | 1 | [[6.48293963e+01 1.70603675e-01] [3.15170604e+02 8.29396325e-01]] |
| **14** | Control | B.1.620 | 0.089571 | 0.764724 | 1 | [[ 64.65879265 0.34120735] [314.34120735 1.65879265]] |
| **15** | Control | P.1 | 0.599516 | 0.438763 | 1 | [[ 64.14698163 0.85301837] [311.85301837 4.14698163]] |
| **16** | Vax | B.1.1.486 | 0.768827 | 0.38058 | 1 | [[3.15170604e+02 8.29396325e-01] [6.48293963e+01 1.70603675e-01]] |
| **17** | Vax | B.1.1.7 | 2.337644 | 0.12628 | 1 | [[ 76.30446194 239.69553806] [ 15.69553806 49.30446194]] |
| **18** | Vax | B.1.2 | 0.271578 | 0.602275 | 1 | [[311.02362205 4.97637795] [ 63.97637795 1.02362205]] |
| **19** | Vax | B.1.214.2 | 0.768827 | 0.38058 | 1 | [[3.15170604e+02 8.29396325e-01] [6.48293963e+01 1.70603675e-01]] |
| **20** | Vax | B.1.234 | 0.768827 | 0.38058 | 1 | [[3.15170604e+02 8.29396325e-01] [6.48293963e+01 1.70603675e-01]] |
| **21** | Vax | B.1.243 | 0.000331 | 0.985479 | 1 | [[313.51181102 2.48818898] [ 64.48818898 0.51181102]] |
| **22** | Vax | B.1.311 | 0.768827 | 0.38058 | 1 | [[3.15170604e+02 8.29396325e-01] [6.48293963e+01 1.70603675e-01]] |
| **23** | Vax | B.1.351 | 0.096163 | 0.756484 | 1 | [[310.19422572 5.80577428] [ 63.80577428 1.19422572]] |
| **24** | Vax | B.1.427 | 0.000331 | 0.985479 | 1 | [[313.51181102 2.48818898] [ 64.48818898 0.51181102]] |
| **25** | Vax | B.1.525 | 0.000331 | 0.985479 | 1 | [[313.51181102 2.48818898] [ 64.48818898 0.51181102]] |
| **26** | Vax | B.1.526 | 8.084124 | 0.004465 | 1 | [[301.07086614 14.92913386] [ 61.92913386 3.07086614]] |
| **27** | Vax | B.1.526.1 | 1.83817 | 0.175166 | 1 | [[284.48293963 31.51706037] [ 58.51706037 6.48293963]] |
| **28** | Vax | B.1.526.3 | 0.089571 | 0.764724 | 1 | [[314.34120735 1.65879265] [ 64.65879265 0.34120735]] |
| **29** | Vax | B.1.596 | 0.768827 | 0.38058 | 1 | [[3.15170604e+02 8.29396325e-01] [6.48293963e+01 1.70603675e-01]] |

| | | | | | | |
|---|---|---|---|---|---|---|
| **30** | Vax | B.1.620 | 0.089571 | 0.764724 | 1 | [[314.34120735 1.65879265] [ 64.65879265 0.34120735]] |
| **31** | Vax | P.1 | 0.599516 | 0.438763 | 1 | [[311.85301837 4.14698163] [ 64.14698163 0.85301837]] |

Supplemental Table 5

| | mut | c | tal vaccinat | Total Contro | p value | dof | expected |
|---|---|---|---|---|---|---|---|
| **0** | S:L5X | 7.364885 | 5 | 4 | 0.006651 | 1 | [[327.5 65.5] [ 7.5 1.5]] |
| **1** | S:L5F | 9.65119 | 10 | 14 | 0.001892 | 1 | [[315. 63.] [ 20. 4.]] |
| **2** | S:S13I | 0.050503 | 0 | 4 | 0.822191 | 1 | [[331.66666667 66.33333333] [ 3.33333333 0.66666667]] |
| **3** | S:L18F | 0.154615 | 3 | 9 | 0.694163 | 1 | [[325. 65.] [ 10. 2.]] |
| **4** | S:L18R | 0.1005 | 0 | 2 | 0.75123 | 1 | [[333.33333333 66.66666667] [ 1.66666667 0.33333333]] |
| **5** | S:T19R | 0.1005 | 0 | 2 | 0.75123 | 1 | [[333.33333333 66.66666667] [ 1.66666667 0.33333333]] |
| **6** | S:T20N | 0.64806 | 2 | 3 | 0.420807 | 1 | [[330.83333333 66.16666667] [ 4.16666667 0.83333333]] |
| **7** | S:T22I | 0.801995 | 0 | 1 | 0.370498 | 1 | [[3.34166667e+02 6.68333333e+01] [8.33333333e-01 1.66666667e- |
| **8** | S:Q23R | 0.1005 | 1 | 1 | 0.75123 | 1 | [[333.33333333 66.66666667] [ 1.66666667 0.33333333]] |
| **9** | S:P26S | 1.860976 | 3 | 4 | 0.172512 | 1 | [[329.16666667 65.83333333] [ 5.83333333 1.16666667]] |
| **10** | S:A27V | 0.801995 | 0 | 1 | 0.370498 | 1 | [[3.34166667e+02 6.68333333e+01] [8.33333333e-01 1.66666667e- |
| **11** | S:T29I | 0.801995 | 0 | 1 | 0.370498 | 1 | [[3.34166667e+02 6.68333333e+01] [8.33333333e-01 1.66666667e- |
| **12** | S:H49Y | 0.162015 | 0 | 5 | 0.687308 | 1 | [[330.83333333 66.16666667] [ 4.16666667 0.83333333]] |
| **13** | S:A67S | 0.1005 | 1 | 1 | 0.75123 | 1 | [[333.33333333 66.66666667] [ 1.66666667 0.33333333]] |
| **14** | S:A67V | 0 | 0 | 3 | 1 | 1 | [[332.5 66.5] [ 2.5 0.5]] |
| **15** | S:H69Y | 0.801995 | 0 | 1 | 0.370498 | 1 | [[3.34166667e+02 6.68333333e+01] [8.33333333e-01 1.66666667e- |
| **16** | S:H69- | 1.355546 | 45 | 251 | 0.244311 | 1 | [[ 88.33333333 17.66666667] [246.66666667 49.33333333]] |
| **17** | S:V70- | 1.355546 | 45 | 251 | 0.244311 | 1 | [[ 88.33333333 17.66666667] [246.66666667 49.33333333]] |
| **18** | S:G75V | 0.801995 | 0 | 1 | 0.370498 | 1 | [[3.34166667e+02 6.68333333e+01] [8.33333333e-01 1.66666667e- |
| **19** | S:T76I | 0.801995 | 0 | 1 | 0.370498 | 1 | [[3.34166667e+02 6.68333333e+01] [8.33333333e-01 1.66666667e- |
| **20** | S:R78M | 0.1005 | 1 | 1 | 0.75123 | 1 | [[333.33333333 66.66666667] [ 1.66666667 0.33333333]] |
| **21** | S:F79L | 0.1005 | 1 | 1 | 0.75123 | 1 | [[333.33333333 66.66666667] [ 1.66666667 0.33333333]] |
| **22** | S:D80A | 0.116311 | 2 | 5 | 0.73307 | 1 | [[329.16666667 65.83333333] [ 5.83333333 1.16666667]] |
| **23** | S:D80G | 0.63769 | 0 | 8 | 0.424548 | 1 | [[328.33333333 65.66666667] [ 6.66666667 1.33333333]] |
| **24** | S:T95I | 10.95774 | 12 | 18 | 0.000932 | 1 | [[310. 62.] [ 25. 5.]] |
| **25** | S:S98F | 0.050503 | 0 | 4 | 0.822191 | 1 | [[331.66666667 66.33333333] [ 3.33333333 0.66666667]] |
| **26** | S:V126A | 0.1005 | 1 | 1 | 0.75123 | 1 | [[333.33333333 66.66666667] [ 1.66666667 0.33333333]] |
| **27** | S:D138Y | 0.116311 | 2 | 5 | 0.73307 | 1 | [[329.16666667 65.83333333] [ 5.83333333 1.16666667]] |
| **28** | S:D138H | 0.050503 | 0 | 4 | 0.822191 | 1 | [[331.66666667 66.33333333] [ 3.33333333 0.66666667]] |
| **29** | S:V143F | 0.801995 | 0 | 1 | 0.370498 | 1 | [[3.34166667e+02 6.68333333e+01] [8.33333333e-01 1.66666667e- |

| | | | | | | | |
|---|---|---|---|---|---|---|---|
| **30** | S:Y144- | 2.782816 | 45 | 260 | 0.095281 | 1 | [[ 80.83333333 16.16666667] [254.16666667 50.83333333]] |
| **31** | S:W152C | 0.050503 | 0 | 4 | 0.822191 | 1 | [[331.66666667 66.33333333] [ 3.33333333 0.66666667]] |
| **32** | S:W152L | 0 | 0 | 3 | 1 | 1 | [[332.5 66.5] [ 2.5 0.5]] |
| **33** | S:S155R | 1.789618 | 2 | 29 | 0.180973 | 1 | [[309.16666667 61.83333333] [ 25.83333333 5.16666667]] |
| **34** | S:E156G | 0.1005 | 0 | 2 | 0.75123 | 1 | [[333.33333333 66.66666667] [ 1.66666667 0.33333333]] |
| **35** | S:F157S | 2.690984 | 2 | 34 | 0.100918 | 1 | [[305. 61.] [ 30. 6.]] |
| **36** | S:F157L | 0.801995 | 0 | 1 | 0.370498 | 1 | [[3.34166667e+02 6.68333333e+01] [8.33333333e-01 1.66666667e- |
| **37** | S:F157- | 0.1005 | 0 | 2 | 0.75123 | 1 | [[333.33333333 66.66666667] [ 1.66666667 0.33333333]] |
| **38** | S:R158S | 0.1005 | 0 | 2 | 0.75123 | 1 | [[333.33333333 66.66666667] [ 1.66666667 0.33333333]] |
| **39** | S:R158- | 0.1005 | 0 | 2 | 0.75123 | 1 | [[333.33333333 66.66666667] [ 1.66666667 0.33333333]] |
| **40** | S:D178Y | 0 | 1 | 8 | 1 | 1 | [[327.5 65.5] [ 7.5 1.5]] |
| **41** | S:G181R | 0.1005 | 1 | 1 | 0.75123 | 1 | [[333.33333333 66.66666667] [ 1.66666667 0.33333333]] |
| **42** | S:R190S | 0 | 1 | 2 | 1 | 1 | [[332.5 66.5] [ 2.5 0.5]] |
| **43** | S:D215G | 0.116311 | 2 | 5 | 0.73307 | 1 | [[329.16666667 65.83333333] [ 5.83333333 1.16666667]] |
| **44** | S:D215H | 0.801995 | 0 | 1 | 0.370498 | 1 | [[3.34166667e+02 6.68333333e+01] [8.33333333e-01 1.66666667e- |
| **45** | S:S221L | 0.050503 | 1 | 3 | 0.822191 | 1 | [[331.66666667 66.33333333] [ 3.33333333 0.66666667]] |
| **46** | S:A222V | 0 | 0 | 3 | 1 | 1 | [[332.5 66.5] [ 2.5 0.5]] |
| **47** | S:I235V | 0.801995 | 0 | 1 | 0.370498 | 1 | [[3.34166667e+02 6.68333333e+01] [8.33333333e-01 1.66666667e- |
| **48** | S:L242H | 0.116311 | 2 | 5 | 0.73307 | 1 | [[329.16666667 65.83333333] [ 5.83333333 1.16666667]] |
| **49** | S:L242- | 0.050503 | 1 | 3 | 0.822191 | 1 | [[331.66666667 66.33333333] [ 3.33333333 0.66666667]] |
| **50** | S:A243- | 0.299093 | 3 | 8 | 0.584451 | 1 | [[325.83333333 65.16666667] [ 9.16666667 1.83333333]] |
| **51** | S:L244X | 0.1005 | 1 | 1 | 0.75123 | 1 | [[333.33333333 66.66666667] [ 1.66666667 0.33333333]] |
| **52** | S:L244- | 0 | 2 | 7 | 1 | 1 | [[327.5 65.5] [ 7.5 1.5]] |
| **53** | S:H245X | 0.1005 | 1 | 1 | 0.75123 | 1 | [[333.33333333 66.66666667] [ 1.66666667 0.33333333]] |
| **54** | S:H245- | 0 | 2 | 7 | 1 | 1 | [[327.5 65.5] [ 7.5 1.5]] |
| **55** | S:R246- | 0.1005 | 0 | 2 | 0.75123 | 1 | [[333.33333333 66.66666667] [ 1.66666667 0.33333333]] |
| **56** | S:D253G | 9.04387 | 9 | 12 | 0.002636 | 1 | [[317.5 63.5] [ 17.5 3.5]] |
| **57** | S:S254F | 0.801995 | 0 | 1 | 0.370498 | 1 | [[3.34166667e+02 6.68333333e+01] [8.33333333e-01 1.66666667e- |
| **58** | S:S255F | 0.801995 | 0 | 1 | 0.370498 | 1 | [[3.34166667e+02 6.68333333e+01] [8.33333333e-01 1.66666667e- |
| **59** | S:W258L | 0.1005 | 0 | 2 | 0.75123 | 1 | [[333.33333333 66.66666667] [ 1.66666667 0.33333333]] |
| **60** | S:T323I | 0.801995 | 0 | 1 | 0.370498 | 1 | [[3.34166667e+02 6.68333333e+01] [8.33333333e-01 1.66666667e- |
| **61** | S:V327I | 0.162015 | 0 | 5 | 0.687308 | 1 | [[330.83333333 66.16666667] [ 4.16666667 0.83333333]] |

| | | | | | | | |
|---|---|---|---|---|---|---|---|
| **62** | S:V367F | 0.1005 | 0 | 2 | 0.75123 | 1 | [[333.33333333 66.66666667] [ 1.66666667 0.33333333]] |
| **63** | S:P384L | 0.1005 | 1 | 1 | 0.75123 | 1 | [[333.33333333 66.66666667] [ 1.66666667 0.33333333]] |
| **64** | S:K417N | 0.116311 | 2 | 5 | 0.73307 | 1 | [[329.16666667 65.83333333] [ 5.83333333 1.16666667]] |
| **65** | S:K417T | 0.64806 | 2 | 3 | 0.420807 | 1 | [[330.83333333 66.16666667] [ 4.16666667 0.83333333]] |
| **66** | S:D427G | 0.801995 | 0 | 1 | 0.370498 | 1 | [[3.34166667e+02 6.68333333e+01] [8.33333333e-01 1.66666667e- |
| **67** | S:N440K | 0.1005 | 0 | 2 | 0.75123 | 1 | [[333.33333333 66.66666667] [ 1.66666667 0.33333333]] |
| **68** | S:L452R | 2.881422 | 2 | 35 | 0.089607 | 1 | [[304.16666667 60.83333333] [ 30.83333333 6.16666667]] |
| **69** | S:L452Q | 0.801995 | 0 | 1 | 0.370498 | 1 | [[3.34166667e+02 6.68333333e+01] [8.33333333e-01 1.66666667e- |
| **70** | S:L452M | 0.801995 | 0 | 1 | 0.370498 | 1 | [[3.34166667e+02 6.68333333e+01] [8.33333333e-01 1.66666667e- |
| **71** | S:S477N | 0.050503 | 1 | 3 | 0.822191 | 1 | [[331.66666667 66.33333333] [ 3.33333333 0.66666667]] |
| **72** | S:T478K | 0 | 0 | 3 | 1 | 1 | [[332.5 66.5] [ 2.5 0.5]] |
| **73** | S:E484K | 8.691305 | 14 | 27 | 0.003197 | 1 | [[300.83333333 60.16666667] [ 34.16666667 6.83333333]] |
| **74** | S:F490S | 0.801995 | 0 | 1 | 0.370498 | 1 | [[3.34166667e+02 6.68333333e+01] [8.33333333e-01 1.66666667e- |
| **75** | S:S494P | 0 | 1 | 8 | 1 | 1 | [[327.5 65.5] [ 7.5 1.5]] |
| **76** | S:N501Y | 0.657115 | 42 | 230 | 0.41758 | 1 | [[108.33333333 21.66666667] [226.66666667 45.33333333]] |
| **77** | S:A570D | 1.079934 | 44 | 244 | 0.298712 | 1 | [[ 95. 19.] [240. 48.]] |
| **78** | S:T572I | 0.801995 | 0 | 1 | 0.370498 | 1 | [[3.34166667e+02 6.68333333e+01] [8.33333333e-01 1.66666667e- |
| **79** | S:D574Y | 0.801995 | 0 | 1 | 0.370498 | 1 | [[3.34166667e+02 6.68333333e+01] [8.33333333e-01 1.66666667e- |
| **80** | S:Q613H | 0.801995 | 0 | 1 | 0.370498 | 1 | [[3.34166667e+02 6.68333333e+01] [8.33333333e-01 1.66666667e- |
| **81** | S:D614G | 0.801995 | 67 | 334 | 0.370498 | 1 | [[8.33333333e-01 1.66666667e-01] [3.34166667e+02 6.68333333e+ |
| **82** | S:A623S | 0.801995 | 0 | 1 | 0.370498 | 1 | [[3.34166667e+02 6.68333333e+01] [8.33333333e-01 1.66666667e- |
| **83** | S:H655Y | 0.64806 | 2 | 3 | 0.420807 | 1 | [[330.83333333 66.16666667] [ 4.16666667 0.83333333]] |
| **84** | S:Q677H | 0.304545 | 2 | 4 | 0.581047 | 1 | [[330. 66.] [ 5. 1.]] |
| **85** | S:N679K | 0.801995 | 0 | 1 | 0.370498 | 1 | [[3.34166667e+02 6.68333333e+01] [8.33333333e-01 1.66666667e- |
| **86** | S:P681H | 1.044258 | 46 | 253 | 0.306833 | 1 | [[ 85.83333333 17.16666667] [249.16666667 49.83333333]] |
| **87** | S:P681R | 0.050503 | 0 | 4 | 0.822191 | 1 | [[331.66666667 66.33333333] [ 3.33333333 0.66666667]] |
| **88** | S:A688V | 0.1005 | 0 | 2 | 0.75123 | 1 | [[333.33333333 66.66666667] [ 1.66666667 0.33333333]] |
| **89** | S:A701V | 10.80182 | 13 | 21 | 0.001014 | 1 | [[306.66666667 61.33333333] [ 28.33333333 5.66666667]] |
| **90** | S:E702D | 0.801995 | 0 | 1 | 0.370498 | 1 | [[3.34166667e+02 6.68333333e+01] [8.33333333e-01 1.66666667e- |
| **91** | S:T716I | 0.423026 | 46 | 246 | 0.515431 | 1 | [[ 91.66666667 18.33333333] [243.33333333 48.66666667]] |
| **92** | S:T732A | 0.1005 | 0 | 2 | 0.75123 | 1 | [[333.33333333 66.66666667] [ 1.66666667 0.33333333]] |
| **93** | S:S735A | 0.1005 | 0 | 2 | 0.75123 | 1 | [[333.33333333 66.66666667] [ 1.66666667 0.33333333]] |

| | | | | | | | |
|---|---|---|---|---|---|---|---|
| **94** | S:G769V | 0 | 0 | 3 | 1 | 1 | [[332.5 66.5] [ 2.5 0.5]] |
| **95** | S:T791I | 0.1005 | 0 | 2 | 0.75123 | 1 | [[333.33333333 66.66666667] [ 1.66666667 0.33333333]] |
| **96** | S:D796H | 0.801995 | 0 | 1 | 0.370498 | 1 | [[3.34166667e+02 6.68333333e+01] [8.33333333e-01 1.66666667e- |
| **97** | S:P812L | 0.801995 | 0 | 1 | 0.370498 | 1 | [[3.34166667e+02 6.68333333e+01] [8.33333333e-01 1.66666667e- |
| **98** | S:P812S | 0.801995 | 0 | 1 | 0.370498 | 1 | [[3.34166667e+02 6.68333333e+01] [8.33333333e-01 1.66666667e- |
| **99** | S:A845S | 0.801995 | 0 | 1 | 0.370498 | 1 | [[3.34166667e+02 6.68333333e+01] [8.33333333e-01 1.66666667e- |
| **100** | S:A846S | 0.801995 | 0 | 1 | 0.370498 | 1 | [[3.34166667e+02 6.68333333e+01] [8.33333333e-01 1.66666667e- |
| **101** | S:A846V | 0.801995 | 0 | 1 | 0.370498 | 1 | [[3.34166667e+02 6.68333333e+01] [8.33333333e-01 1.66666667e- |
| **102** | S:T859N | 2.881422 | 2 | 35 | 0.089607 | 1 | [[304.16666667 60.83333333] [ 30.83333333 6.16666667]] |
| **103** | S:F888L | 0 | 0 | 3 | 1 | 1 | [[332.5 66.5] [ 2.5 0.5]] |
| **104** | S:D936N | 0.801995 | 0 | 1 | 0.370498 | 1 | [[3.34166667e+02 6.68333333e+01] [8.33333333e-01 1.66666667e- |
| **105** | S:S939F | 0.1005 | 0 | 2 | 0.75123 | 1 | [[333.33333333 66.66666667] [ 1.66666667 0.33333333]] |
| **106** | S:D950H | 2.690984 | 2 | 34 | 0.100918 | 1 | [[305. 61.] [ 30. 6.]] |
| **107** | S:D950N | 0.801995 | 0 | 1 | 0.370498 | 1 | [[3.34166667e+02 6.68333333e+01] [8.33333333e-01 1.66666667e- |
| **108** | S:Q957R | 0.801995 | 0 | 1 | 0.370498 | 1 | [[3.34166667e+02 6.68333333e+01] [8.33333333e-01 1.66666667e- |
| **109** | S:T961M | 0.801995 | 0 | 1 | 0.370498 | 1 | [[3.34166667e+02 6.68333333e+01] [8.33333333e-01 1.66666667e- |
| **110** | S:S982A | 1.079934 | 44 | 244 | 0.298712 | 1 | [[ 95. 19.] [240. 48.]] |
| **111** | S:A1020S | 0.801995 | 0 | 1 | 0.370498 | 1 | [[3.34166667e+02 6.68333333e+01] [8.33333333e-01 1.66666667e- |
| **112** | S:T1027I | 0.818321 | 3 | 6 | 0.365672 | 1 | [[327.5 65.5] [ 7.5 1.5]] |
| **113** | S:V1060I | 0.801995 | 0 | 1 | 0.370498 | 1 | [[3.34166667e+02 6.68333333e+01] [8.33333333e-01 1.66666667e- |
| **114** | S:A1087S | 0.801995 | 0 | 1 | 0.370498 | 1 | [[3.34166667e+02 6.68333333e+01] [8.33333333e-01 1.66666667e- |
| **115** | S:H1101Q | 0.801995 | 0 | 1 | 0.370498 | 1 | [[3.34166667e+02 6.68333333e+01] [8.33333333e-01 1.66666667e- |
| **116** | S:D1118H | 0.903624 | 45 | 247 | 0.341812 | 1 | [[ 91.66666667 18.33333333] [243.33333333 48.66666667]] |
| **117** | S:G1167V | 0.801995 | 0 | 1 | 0.370498 | 1 | [[3.34166667e+02 6.68333333e+01] [8.33333333e-01 1.66666667e- |
| **118** | S:D1168G | 0.801995 | 0 | 1 | 0.370498 | 1 | [[3.34166667e+02 6.68333333e+01] [8.33333333e-01 1.66666667e- |
| **119** | S:V1176F | 0.64806 | 2 | 3 | 0.420807 | 1 | [[330.83333333 66.16666667] [ 4.16666667 0.83333333]] |
| **120** | S:K1191N | 0 | 2 | 13 | 1 | 1 | [[322.5 64.5] [ 12.5 2.5]] |
| **121** | S:G1219V | 1.005 | 0 | 10 | 0.316104 | 1 | [[326.66666667 65.33333333] [ 8.33333333 1.66666667]] |
| **122** | S:M1237I | 0.801995 | 0 | 1 | 0.370498 | 1 | [[3.34166667e+02 6.68333333e+01] [8.33333333e-01 1.66666667e- |
| **123** | S:C1243F | 0.801995 | 0 | 1 | 0.370498 | 1 | [[3.34166667e+02 6.68333333e+01] [8.33333333e-01 1.66666667e- |
| **124** | S:V1264L | 0.1005 | 1 | 1 | 0.75123 | 1 | [[333.33333333 66.66666667] [ 1.66666667 0.33333333]] |
| **125** | S:K1266N | 0.801995 | 0 | 1 | 0.370498 | 1 | [[3.34166667e+02 6.68333333e+01] [8.33333333e-01 1.66666667e- |

01]]

01]]
01]]

01]]

01]]
01]]

01]]

01]]

01]]

01]]

01]]
01]]

01]]

01]]

01]]
01]]

01]]

01]]
01]]
01]]
01]]
01]]

01]]

01]]

01]]
01]]
01]]
01]]
01]]
01]]

01]]

01]]
01]]
01]]

01]]

01]]
01]]
01]]

01]]
01]]

01]]
01]]

01]]
